# Design, conduct, analysis, and reporting of therapeutic efficacy studies in Visceral Leishmaniasis: A systematic review of published reports, 2000-2021

**DOI:** 10.1101/2023.09.06.23295148

**Authors:** Prabin Dahal, Sauman Singh-Phulgenda, Caitlin Naylor, Matthew Brack, Mitali Chatterjee, Fabiana Alves, Philippe J Guerin, Kasia Stepniewska

## Abstract

A systematic review (SR) of published efficacy studies in visceral leishmaniasis (VL) was carried out to describe methodological aspect of design, analysis, conduct and reporting. Studies published during 2000-2021 and indexed in the Infectious Diseases Data Observatory (IDDO) VL library of clinical studies were eligible for inclusion (n=89 studies). The IDDO VL library is a living SR of prospective therapeutic studies (PROSPERO: CRD42021284622) and is updated bi-annually. A total of 40 (44.9%) studies were randomised, 33 (37.1%) were single-armed, 14 (15.7%) were non-randomised multi-armed studies, and randomisation was unclear in 2 (2.2%). After initial screening, patients were enrolled into the study upon confirmation of VL using parasitological method in 26 (29.2%), and serological and parasitological method in 63 (70.8%). Post-treatment follow-up duration was <6months in 3 (3.3%) studies, 6-months in 75 (84.3%), and >6months in 11 (12.4%) studies. Relapse was defined solely based on clinical suspicion in 4 (4.5%) studies, parasitological demonstration in 64 (71.9%), using molecular/serological/parasitological in 6 (6.7%), and was unclear in 15 (16.9%). Quality control of laboratory measures adopted was unclear in 66 (74.2%) studies, sample size calculation was reported in only 34 (38.2%) studies, and cured proportion was presented only as a point estimate in 39 (43.8%) studies. This review highlights substantial variations in definitions adopted for patient screening, disease diagnosis and therapeutic outcomes suggesting an urgent need for harmonisation of VL clinical trials protocol.

## Introduction

The first randomised trial for comparing efficacy of treatment regimens in visceral leishmaniasis (VL) was conducted in 1983 ^1^. Since then several randomised and non-randomised studies have evaluated the therapeutic efficacy of antileishmanial drugs ^2^. Previous reviews have characterised the spectrum of patient characteristics, treatment regimens adopted, and completeness of reporting of clinical and safety outcomes in VL therapeutic studies ^2,3^. A thorough review on methodological aspects of design, conduct, analysis, and reporting of VL clinical therapeutic efficacy studies is currently lacking. Such review has been carried out in the context of cutaneous leishmaniasis ^4–7^, leading to preparation of a guidance document on optimal approaches for design, conduct, analysis and reporting of CL studies ^7^.

This review was conducted with an overall aim to characterise different aspects of study design, conduct, analysis and reporting of clinical therapeutic efficacy studies in VL published since 2000.

## Material and methods

### Information sources and search strategy

This review synthesises data from studies indexed in the Infectious Diseases Data Observatory (IDDO) VL library of clinical studies ^8^. The IDDO library indexes any prospective clinical studies describing efficacy of any antileishmanial therapies published since 1980; details on the search strategy adopted for each of the databases is described elsewhere ^2^. Briefly, the IDDO VL library is a living systematic (VL LSR) updated bi-annually and searches the following databases: Ovid Embase, Scopus, Web of Science Core Collection, Cochrane Central Register of Controlled Trials, clinicaltrials.gov, WHO ICTRP, as well as IMEMR, IMSEAR, and LILACS from the WHO Global Index Medicus. All the studies indexed in the IDDO living systematic review was eligible for inclusion in this review.

### Study selection and data extraction

For the purpose of this review, studies published on or after 2000 was considered. Data on the following aspects of design and conduct of studies captured by the IDDO LSR were extracted: inclusion & exclusion criteria, case definition used, sample source used for parasitological confirmation of the disease, randomisation, blinding, follow-up duration, the number of participants enrolled, endpoints adopted and their definitions, and details of the laboratory procedures adopted.

### Data summary

Descriptive summaries were presented for the characteristics of the studies included in the review. No patient related outcome data were analysed in this review and hence risk of bias assessment in studies included was not carried out.

## Results

The IDDO living systematic review has indexed 89 studies published from 01/01/2000 through to 17/11/2021 ^8^. There were 61 (68.5%) studies from the Indian subcontinent, 16 (18.0%) from East Africa, 4 (4.5%) from the Mediterranean region, 4 (4.5%) from South America, 3 (3.4%) from Central Asia (the Middle East), and 1 (1.1%) multi-regional study. Of the 89 studies, 28(31.5%) studies were published during 2000-2004, 20 (22.5%) during 2005-2009, 23(25.8%) during 2010-2014, and 18 (20.2%) were published on or after 2015. Overall, there were 27,070 patients enrolled in 187 drug arms with a median sample size of 51 (range: 1–3,126) patients per arm. Further description of the studies included is presented in supplemental file (S1).

### Study design and conduct

A total of 40 (44.9%) studies were randomised, 33 (37.1%) were single-armed studies, 14 (15.7%) were non-randomised multi-armed studies, and randomisation status was unclear in 2 (2.2%). Of the 89 trials, 59 (66.3%) were open-label studies, 2 (2.2%) were blinded, and the blinding status was not stated in the remaining 28 (31.5%) studies. The median sample size per study for RCTs was 152 [Interquartile range (IQR): 84-400; range: 25-1,485] and non-randomised studies were 120 [IQR: 60-309; 12-3,126].

### Randomised studies (n=40)

Block randomisation was used in 12 studies (of which 3 used permuted block randomisation); block sizes ranged from 4-28 (**Figure 1**). Randomisation was carried out by balancing the treatment regimens on at least one prognostic factor in 3 studies; while the randomisation details were unclear in 25 studies (**Figure 1** and **supplemental file (S1))**. Sequence generation was carried out using a computerised system in 22 (55.0%), using a random number table in 2 (5.0%), and the methodology was unclear in 16 (40.0%) (**Figure 1**). Treatment allocation was concealed using a sealed, opaque envelope/box in 22 (55.0%) studies and allocation concealment was unclear in 18 (45.0%) studies (**Figure 1**). Of the 40 randomised studies, 2 were blinded, 33 were open labelled, and blinding status was unclear in 5 studies.

**Figure 1:** Details of randomisation method and allocation concealment included in the randomised studies included in the review.

### Inclusion and exclusion criteria adopted

A complete list of inclusion and exclusion criteria adopted for patient enrolment is presented in **supplemental file (S2)**.

### Informed consent

Of the 89 studies, requirement of informed consent (or assent for children) was stated as a part of inclusion/exclusion criteria in 21 (23.6%) studies, wasn’t explicitly stated as a part of the inclusion/exclusion criteria but was collected before patient enrolment in 57 (64.0%) studies and there was no statement regarding this in the remaining 11 (12.4%) studies (**Table 2**).

### Eligible age-range and included age-range

Eligible age-range for inclusion was children less than 15 years in 12 (13.5%) studies, adults (≥18 years) in 6 (6.7%), patient of all ages in 53 (59.6%) and age-range was not defined as a part of inclusion/exclusion criteria in the remaining 18 (20.2%) studies (**Table 1**).

**Table 1:**
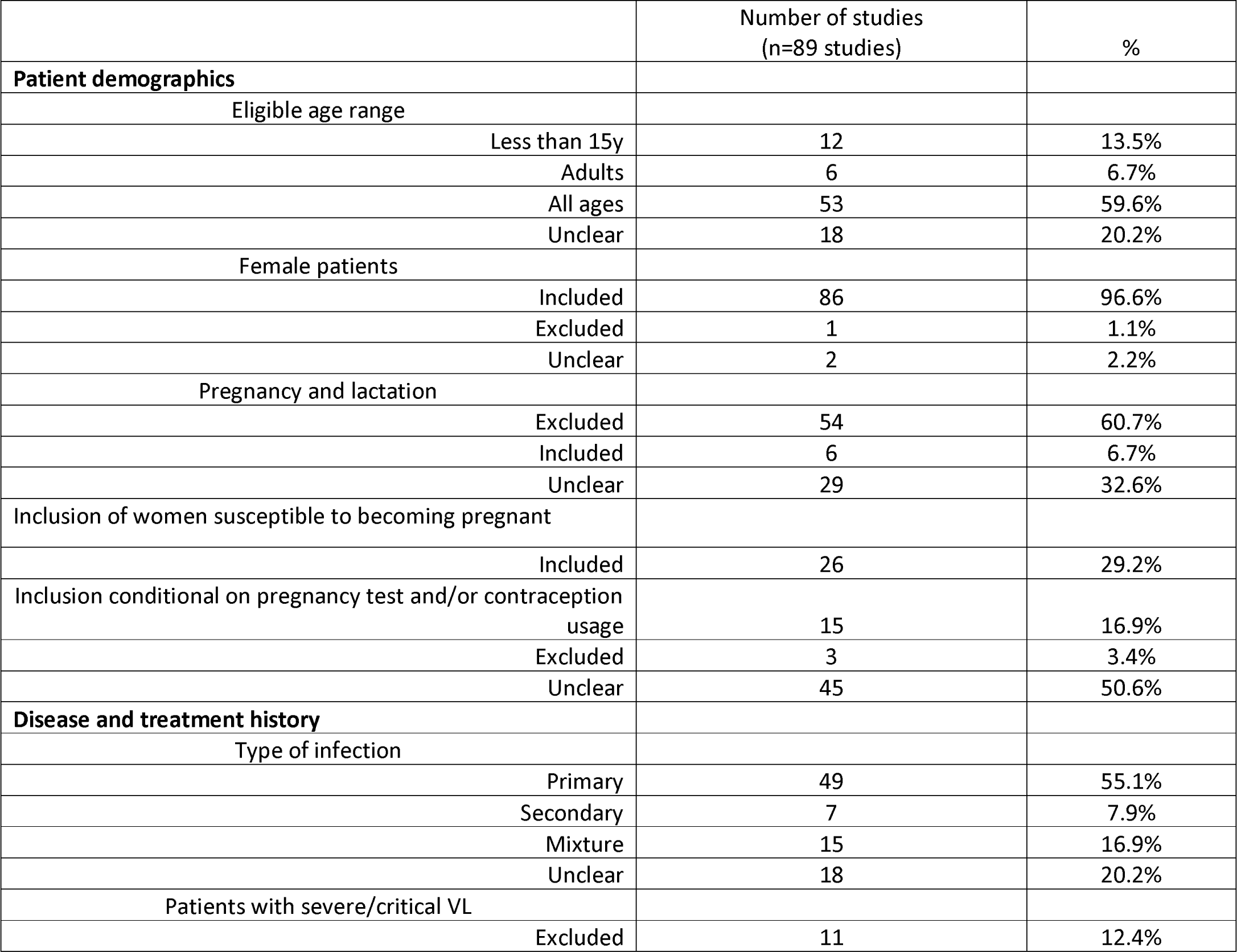
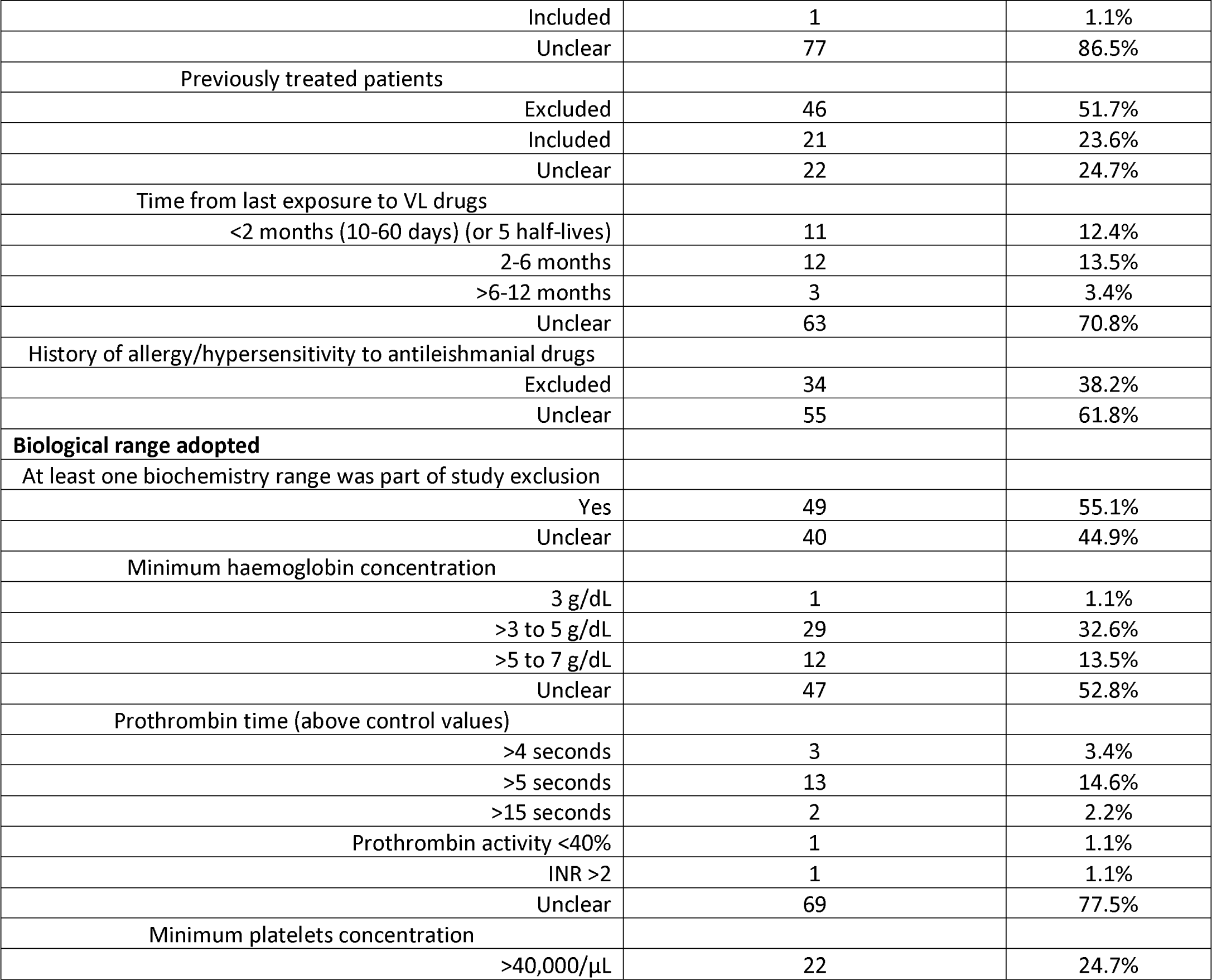
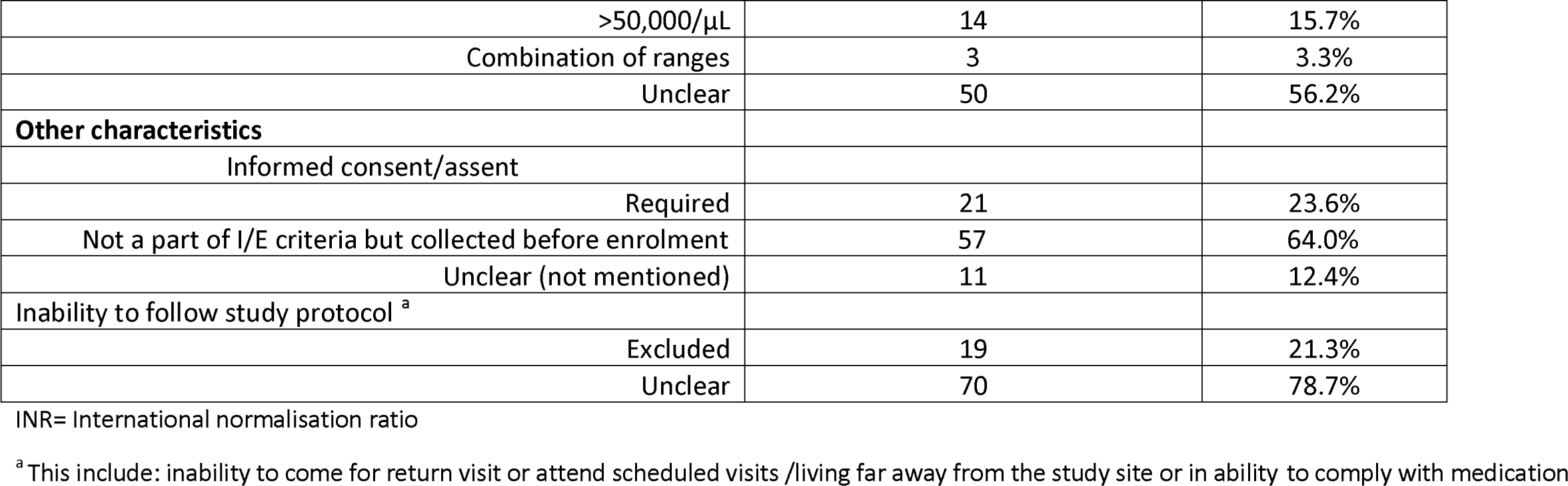
Demographic, disease and biological range adopted for defining inclusion/exclusion of patients.

The age-range of included patients were: children less than 15 years in 13 (14.6%), adults in 7 (7.9%), and patients of all ages in 69 (77.5%) studies. Infants were included in 5 (5.6%) studies, excluded in 72 (80.9%) and their inclusion was unclear in the remaining 12 (13.5%) studies. The maximum eligible/included age was 65 years in 16 (18.0%) studies, the upper range of was 66-80 years in 10 (11.2%) studies and the age-range was unclear in 18 (20.2%) studies (**supplemental file S2).**

### Enrolment of female, pregnant women, and women susceptible to becoming pregnant

Females were excluded in 1 (1.1%) study, included in 86 (96.6%) studies and their inclusion/exclusion was not clear in the remaining 2 (2.2%) studies. Pregnant and lactating women were excluded in 54 (60.7%) studies, included in 6 (6.7%) and their inclusion/exclusion couldn’t be discerned in the remaining 29 (32.6%) studies (**Table 1 and Supplemental file S2**).

Women of child bearing age (or those who had reached menarche) were excluded in 3 (3.4%) studies, their inclusion was conditional on negative pregnancy test or willingness to undertake contraception in 15 (16.9%) studies, they were included in 26 (29.2%) studies without description of pregnancy test/contraception usage requirements, their inclusion was not clear in the remaining 45 (50.6%) studies.

### Co-morbidities

Patients with HIV, tuberculosis, hepatic, renal and cardiac disorders were the most common co-morbidities as a part of the exclusion criteria adopted (**Figure 2**). Patients with at least one co-morbidity were clearly excluded in 58 (65.2%) studies, included in 7 (7.9%) studies and this couldn’t be discerned in the remaining 24 (27.0%) studies. In particular, patients with HIV co-infections were excluded in 64 (71.9%), included in 11 (12.4%) studies and unclear in the rest 14 (15.7%) studies). Those with hepatic disorders were excluded in 31 (34.8%) and included in 2 (2.2%) studies (unclear in the remaining 56 studies), patients with TB co-infections were excluded in 34 (38.2%) studies and included in 5 (5.6%) (unclear in the remaining 50 studies), patients with renal disorders were excluded in 27 (30.3%) studies, and those with cardiac disorders were excluded in 21 (23.6%) studies. Co-morbidities reported included: malnutrition, helminth co-infections, malaria, endocrine disorders such as diabetes and pancreatitis, hypertension, PKDL, hearing disorders and bleeding disorders (**Figure 2**).

**Figure 2:** Co-morbidities as exclusion criteria and reasons for patient exclusion in the studies included in the review Legend: Panel A: Endocrine disorders included patients with diabetes and pancreatitis; Hepatic disorders including jaundice, hepatitis or hepatic encephalopathy; any pulmonary condition included pneumonia, TB or any respiratory illness; bleeding diathesis including coagulation disorders, G6PD deficiency or any haematological disorders. Panel B: This presents data from 46 studies that clearly reported the patient flow. Panel B presents the sample size by study design on logarithm (base10) scale.

### Malnutrition

Malnourished patients were included in 12 (13.5%) studies, excluded in 12 (13.5%) and the information was not clear in 65 (73.0%) studies. Of the 24 studies that clearly indicated inclusion/exclusion of malnourished patients, 14 studies used anthropometric indicators (BMI/WHZ/WAZ /MUAC/Wasting or some other standards), 2 used clinical definition (kwashiorkor/marasmus/Protein energy malnutrition/Gomez criteria) and the criteria used for assessment of malnutrition was not clear in 6 studies (**Supplemental file S2**).

### Treatment history

Patients with a history of treatment with any antileishmanial treatment within a pre-defined period (time ranged from 10 days to 12 months) were excluded in 46 (51.7%), included in 21 (23.6%) studies and it was unclear in 22 (24.7% studies). Those with a history of hypersensitivity/allergy to the study drug or other antileishmanial therapies were excluded in 34 (38.2%) studies (**Table 1**).

### Parasite related criteria

Patients with freshly diagnosed VL (primary infection) were recruited in 49 (55.1%) studies, patients with secondary cases (including resistant, previously unresponsive or relapsing cases) were recruited in 7 (7.9%) studies, a mixture of primary and secondary infections were included in 15 (16.9%) and this was unclear in 18 (20.2%) studies (**Table 1**).

### Disease severity

Patients who were described as severe or critical visceral leishmaniasis were excluded in 11 (12.4%), included in 1 (1.1%), and this was unclear in the remaining 77 (86.5%) studies (**Table 1**). Of the 12 studies that clearly stated inclusion/exclusion of severe/critical VL, the definition of severe or critical VL was based on clinical signs (such as generalised oedema, jaundice, bleeding) in 2 studies, based on biochemistry ranges in 5 studies, and the definition was not presented in the remaining 5 studies (**Supplemental file S1**).

### Haematological measures

#### Haemoglobin

Minimum haemoglobin concentration required for inclusion in the study was 3 g/dL in 1 (1.1%) studies, between >3-5 g/dL in 29 (32.6%) studies, >5-7 g/dL in 12 (13.5%) studies and it was unclear in the remaining 47 (52.8%) studies (**Table 1, Figure 2**).

#### Platelets

The exclusion threshold for platelet counts was <40,000/μL in 22 (24.7%) studies, <50,000/μL in 14 (15.7%), range of other thresholds were used in 3 (3.3%) studies and the eligible platelets range was not reported in the remaining 50 (56.2%) studies (**Table 2, Figure 2**).

**Table 2:** Case definition used for patient screening and confirmation of disease status.

| Case definition for patient screening | Number of studies<br>(n=89 studies) | % |
| --- | --- | --- |
| Compatible clinical diagnosis <sup>a</sup><br>(fever, splenomegaly etc.) | 77 | 86.5% |
| Not specified<br>(no defined criteria stated) | 12 | 13.5% |
| **Definition of compatible clinical diagnosis** |  |  |
| Fever or splenomegaly or cytopenia | 3 | 3.4% |
| Fever + splenomegaly | 18 | 20.2% |
| Fever + hepatomegaly/splenomegaly | 3 | 3.40% |
| Fever + splenomegaly/wasting + weight loss/loss of appetite | 19 | 21.3% |
| fever + splenomegaly + haematological measurement | 6 | 6.6% |
| Fever + splenomegaly + chills + rigor | 3 | 3.4% |
| Fever + splenomegaly/wasting + weight loss/loss of appetite +<br>haematological measurement | 7 | 7.7% |
| Not specified | 30 | 33.7% |
| **Length of fever used in case definition for patient screening** |  |  |
| >1 week | 1 | 1.1% |
| ≥ 2 weeks | 19 | 21.3% |
| Not specified | 69 | 77.6% |
| **Case confirmation method** |  |  |
| Parasitological | 26 | 29.2% |
| Serological and/or parasitological | 56 | 62.9% |
| Serological + parasitological | 7 | 7.9% |

<sup>a</sup> The following were part of the definition of suspected cases of Visceral leishmaniasis: fever, splenomegaly, hepatomegaly, hepatosplenomegaly, chills, rigor, weight loss, loss of appetite, wasting, epistaxis, anaemia, weakness, asthenia, cytopenia, leukopenia, thrombocytopenia, or lymphadenopathy

#### White blood cells (WBC)

Exclusion threshold of granulocytes <1,000/µl was adopted in 13 (14.6%) studies, WBC counts <750/μL was adopted in 1 (1.1%) study, WBC counts <1,000/μL in 18 (20.2%) studies, WBC <2,000/μL in 4 (4.5%) studies, and WBC threshold was not specified in eligibility criteria in 53 (59.6%) studies (**Supplemental file S1**).

#### Liver enzymes and Liver function

##### Albumin

Serum albumin concentration < 2.0 g/dL was an exclusion criterion in 2 (2.2%) studies, >3 times the upper limit of normal (ULN) were excluded in 3 (3.4%), significant proteinuria in 1 (1.1%), and it was not part of the eligibility criteria in the remaining 83 (93.3%) studies (**Supplemental file S2**).

##### Bilirubin

Exclusion range of bilirubin concentration was: >3 times the upper limit of normal (ULN) in 9 (10.1%) studies, >2 ULN in 7 (7.9%), >1.5 ULN in 2 (2.2%), >ULN in 1 (1.1%), >2 normal range in 3 (3.4%), >5 normal range in 1 (1.1%) study, bilirubin concentration higher than 34.2µmol/L to 221 μmol/L were used in 6 (6.7%) studies, and bilirubin was not part of the eligibility criteria in the remaining 60 (67.4%) studies (**Supplemental file S2**).

##### Aminotransferases

Exclusion range of aminotransferase (ASAT/ALAT) measurements were: >2.5 ULN in 3 (3.4%) studies, >3 ULN in 23 (25.8%) studies, >4 ULN in 2 (2.2%) studies, > 5 ULN in 3 (3.4%), > 10 ULN in 1 (1.1%), >3 times the normal range in 3 (3.4%) studies, >5 times the normal range in 1 (1.1%), >200 IU in 1 (1.1%) and aminotransferase levels were not part of eligibility criteria in 52 (58.4%) studies included in this review (**Supplemental file S2**).

##### Prothrombin time (PT)

PT was clearly reported as a part of exclusion criteria in 20 (22.5%) studies: PT >4 seconds above the control was required in 3 studies, >5 seconds was required in 13 studies, PT >15 seconds in 2 studies, INR ratio >2 was required in 2 studies, and further breakdown in presented in **Table 1**.

#### Renal function

##### Serum creatinine

Exclusion range adopted for creatinine concentration were: >1.5 mg/dL in 1 (1.1%) study, >2.0 mg/dl in 7 (7.9%), outside or above the normal range (without further details) in 10 (11.2%) studies, >1.5 ULN/Normal range in 14 (15.7%) studies, > 2 ULN in 3 (3.4%), >3 ULN in 1 (1.1%), and >1.5 normal limit in 3 (3.4%), and creatinine wasn’t part of the eligibility criteria in the remaining 50 (56.2%) studies (**Supplemental file S2**).

##### Blood urea nitrogen (BUN)

Exclusion range adopted for BUN were: >1.5 ULN/Normal range in 11 (12.4%) studies and BUN measurements were not part of eligibility criteria in the remaining 78 (87.6%) studies.

##### Urine urea concentration

Patients with urine urea concentration > 2 × ULN were excluded in 2 (2.2%) studies and this was not mentioned in the remaining 87 (97.8%) studies.

#### Other characteristics

Ability to comply with the scheduled follow-up or proximity to the study centre was stated as an essential criterion for inclusion in 19 (21.3%) studies. Other occasionally adopted exclusion criteria were: the use of prohibited/contraindicated drugs (n=7 studies), alcohol/drug abuse (n=5 studies), life expectancy of <6 months (n=2 studies), undergoing major surgical procedure (n=2 studies), contraindication for splenic/bone marrow aspirate (n=3 studies), and abnormal potassium concentration (n=1 study) (**Supplemental file S2**).

#### Patient screening

##### Case definition for patient screening

Case definition adopted for patient screening was presented in 77 (86.5%) studies with no information in the remaining 12 (13.5%) studies. Overall, 17 different signs and symptoms were part of the case definition adopted in various combinations (**Table 2**). Case definition adopted constituted a combination of fever and splenomegaly/hepatomegaly in 24 (27.0%) studies, a combination of fever and hepatosplenomegaly and weight loss/loss of appetite in 14 (15.7%) studies, a combination of fever, splenomegaly and a haematological measure (cytopenia/anaemia/thrombocytopaenia) in 6 (6.7%) studies, the definition was unclear in 30 (33.7%) studies and the combination of clinical factors used in the remaining 15 (16.9%) studies is presented in **Table 2**.

##### Disease confirmation method

Patients who satisfied the case definition of VL were enrolled into the study upon confirmation of the disease using parasitological confirmation (demonstration of parasite in a tissue aspirate) in 26 (29.2%) studies, using a combination of serological and/or parasitological in the remaining 63 (70.8%) studies.

##### Reasons for patient exclusion

Of the 89 studies included in this review, 46 (51.7%) studies clearly presented patient flow diagram (or CONSORT checklist) (**See supplemental file S2**). Overall, 22,056 patients were screened in these 46 studies of whom 13,878 (62.9%) patients were enrolled. Of the 8,178 who were excluded, 2723 (33.3%) had negative parasitaemia upon parasitological/serological examination, 687 (8.4%) had biochemistry/biological measurements outside of permissible range, 515 (6.3%) of the patients were outside the inclusion age range, 350 (4.3%) patients refused to participate or give consent, and further breakdown of reasons for exclusion is presented in **Figure 2**.

#### Details of the laboratory procedures adopted for patient enrolment

##### Parasite speciation and **Leishmania** zymodeme (isoenzyme) characterisation

*L. donovani* (LD) was the stated as the causative parasites of VL in 23 (25.8%) studies, *L. infantum / L. chagasi* in 3 (3.4%) studies, and the parasites species (genus *Leishmania*) was not stated in the remaining 63 (70.8%) (VL is caused by LD in East Africa and Indian sub-continent, and *L. infantum* in the Mediterranean region and South America ^9^) (**Supplemental file S2**). Isoenzyme characterisation to study the parasite strains were explicitly carried out in 2 studies; one study was from the Mediterranean region ^10^ and the other in HIV negative patients in Eastern Africa ^11^)(**Table 3**).

**Table 3:**
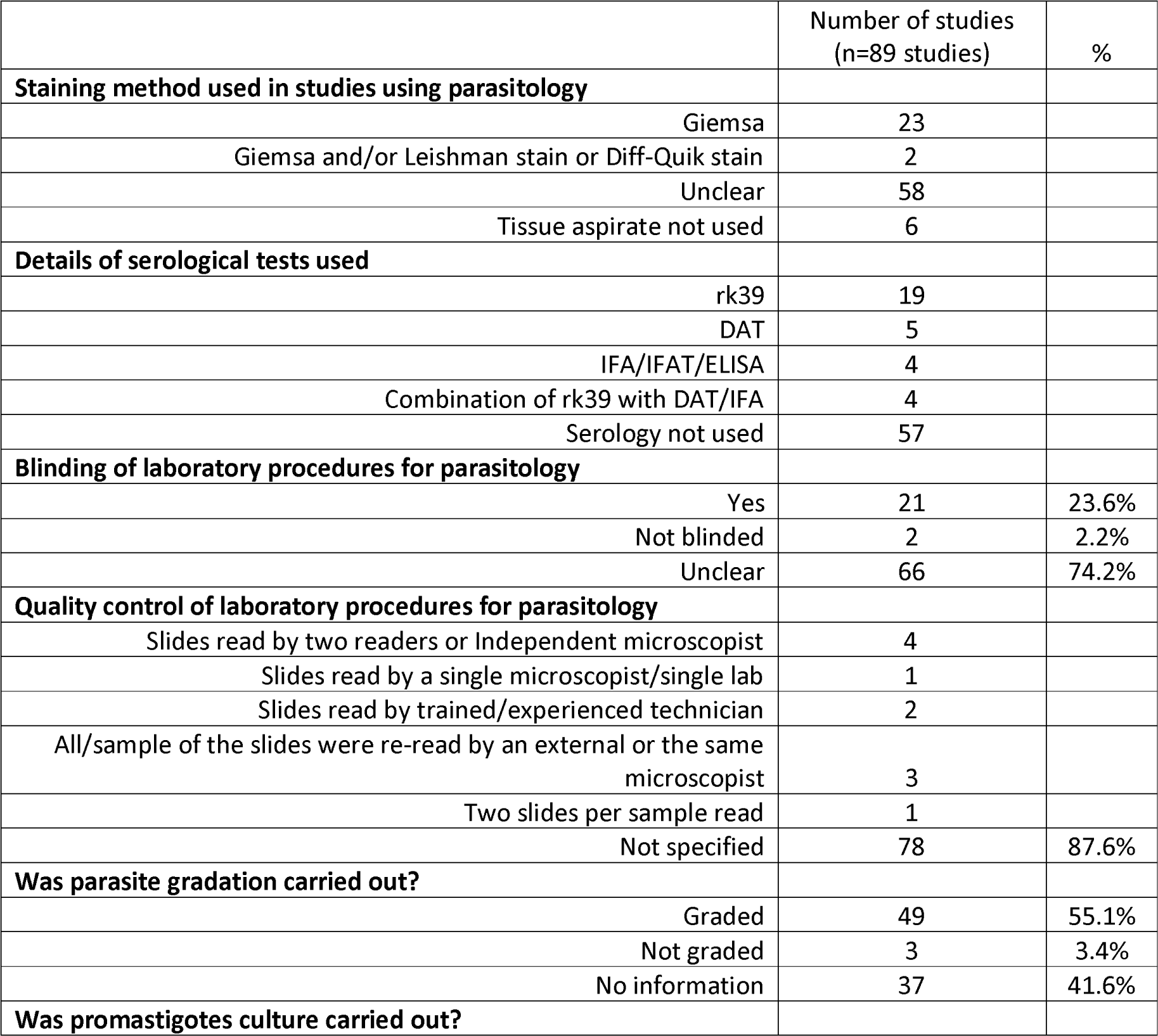
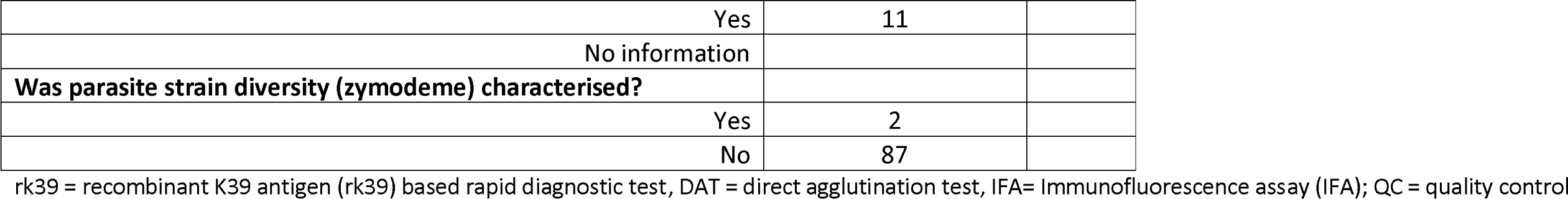
Details of the laboratory procedures adopted.

|  | Number of studies<br>(n=89 studies) | % |
| --- | --- | --- |
| **Staining method used in studies using parasitology** |  |  |
| Giemsa | 23 |  |
| Giemsa and/or Leishman stain or Diff-Quik stain | 2 |  |
| Unclear | 58 |  |
| Tissue aspirate not used | 6 |  |
| **Details of serological tests used** |  |  |
| rk39 | 19 |  |
| DAT | 5 |  |
| IFA/IFAT/ELISA | 4 |  |
| Combination of rk39 with DAT/IFA | 4 |  |
| Serology not used | 57 |  |
| **Blinding of laboratory procedures for parasitology** |  |  |
| Yes | 21 | 23.6% |
| Not blinded | 2 | 2.2% |
| Unclear | 66 | 74.2% |
| **Quality control of laboratory procedures for parasitology** |  |  |
| Slides read by two readers or Independent microscopist | 4 |  |
| Slides read by a single microscopist/single lab | 1 |  |
| Slides read by trained/experienced technician | 2 |  |
| All/sample of the slides were re-read by an external or the same microscopist | 3 |  |
| Two slides per sample read | 1 |  |
| Not specified | 78 | 87.6% |
| **Was parasite gradation carried out?** |  |  |
| Graded | 49 | 55.1% |
| Not graded | 3 | 3.4% |
| No information | 37 | 41.6% |
| **Was promastigotes culture carried out?** |  |  |
| Yes | 11 |  |
| No information |  |  |
| **Was parasite strain diversity (zymodeme) characterised?** |  |  |
| Yes | 2 |  |
| No | 87 |  |

rk39 = recombinant K39 antigen (rk39) based rapid diagnostic test, DAT = direct agglutination test, IFA= Immunofluorescence assay (IFA); QC = quality control

##### Parasite staging (smear and culture)

Parasitological demonstration was based on the identification of amastigotes form of the parasites from tissue smears in 15 (16.9%) studies, and this was not explicitly stated in the remaining 74 (83.1%) studies. In addition, culture (using biphasic medium: Novy– McNeal Nicolle medium) demonstrating the promastigotes form of the parasites were carried out in 11 (12.4%) (**Table 3**).

##### Tissue aspiration

Overall, 83 studies used parasitological method (either alone or in combination of other methods such as serological, culture or molecular) for confirming the presence of parasites and 6 studies used only serological method. Of the 83 studies that used parasitological method, Giemsa stain was used in 23, Giemsa or Leishman stain or Diff-Quik stain was used in 2 studies, and the staining method was not stated in 58 studies. Splenic tissue aspiration was used in 27 (30.3%) studies, a combination of bone marrow and/or spleen or lymph node aspirate in 53 (59.6%), blood sample was used in 4 (4.5%), and the sample used was unclear in 5 (5.6%) (**Supplemental file S2**).

##### Parasite enumeration

Parasitaemia was graded in 49 (55.1%) studies, gradation was not done in 3 (3.4%) studies, and information regarding gradation was not reported in 37 (41.6%) studies (**Table 3**). Parasite enumeration was based on gradation of parasitaemia from tissue aspirates under microscopic fields in 38 (42.7%) studies (using a semi-quantitative scale), and using PCR in 2 (2.2%) studies, parasite gradation was not done in 3 (3.4%), and the information was unclear in 46 (51.7%). Of the 2 studies that adopted PCR method, one study quantified parasitaemia only using peripheral blood ^12^, and the other used both semi-quantitative microscopy counts on tissue aspirate and parasite load in peripheral blood using PCR ^11^ (**Supplemental file S2**).

##### Details of serological method

A total of 32 studies used serological methods for confirmation the presence of the parasites (alone or in combination with other methods). Of these 32 studies, recombinant K39 antigen (rk39) based rapid diagnostic test was used in 19, direct agglutination test (DAT) was used in 5, Immunofluorescence assay (IFA or IFAT or ELISA) in 4 and a combination of rk39 with DAT or IFA in the remaining 4 studies (**Table 3**).

##### Quality control of lab procedures

In 21 (23.6%) studies, the laboratory procedures were blinded to the treatment regimen, 2 (2.2%) studies did not blind the laboratory procedures and this information couldn’t be gathered for the remaining 66 (74.2%) studies. Details regarding any quality control aspect of the laboratory procedures adopted were not clear in 78 (87.6%) studies with only 11 (12.4%) studies reported carrying out quality control of laboratory procedures adopted. Of these 11studies, the slides were read by two readers/independent microscopist in 4 studies, all or a random sample of the slides were re-read by an external or the same microscopist in the 3 studies, two slides were read per sample in 1 study, only read by a single reader in 1 study, and slides were read by trained/experienced technician in 2 studies (**Table 3**).

#### Patient outcome assessments

##### Initial assessment of test of cure (TOC) and tissue aspiration

Initial assessment upon completion of treatment regimen was carried out solely based on clinical assessment in 13 (14.6%) studies, parasitological assessment (with or without clinical assessment) was carried out in 71 (79.8%) studies and the details were not reported in 5 (5.6%) studies. TOC evaluation (either clinical cure, parasitological cure, or clinical and parasitological cure) was carried out within 14 days of treatment initiation in 3 (3.4%) studies, between 15 to 30 days in 68 (76.4%) studies, between 31 to 70 days in 7 (7.9%) studies, mixture of different time-points in 2 (2.2%) studies, and the time of assessment was unclear in 9 (10.1%) (**Table 4**).

**Table 4:** Assessment at end of the treatment and at the end of the study follow-up.

|  | Number of studies<br>(n=89 studies) | % |
| --- | --- | --- |
| **Primary endpoint adopted** |  |  |
| Initial cure or negative parasitaemia at the end of therapy | 12 | 13.5% |
| Ultimate cure/ definitive cure/final cure at end of 6-months or longer | 27 | 30.3% |
| Pharmacokinetics measurements at day 21 | 1 | 1.1% |
| Safety outcomes | 5 | 5.6% |
| Not explicitly stated | 44 | 49.4% |
| **Assessment at end of the treatment (Test of cure assessment) <sup>a</sup>** |  |  |
| Clinical | 13 | 14.6% |
| Clinical + parasitological | 69 | 77.5% |
| Clinical and/or parasitological | 2 | 2.2% |
| No information | 5 | 5.6% |
| **Time point of test of cure assessment** |  |  |
| <15 days | 3 | 3.4% |
| 15 to 30 days | 68 | 76.4% |
| 31 to 70 days | 7 | 7.9% |
| Mixture of one or more of the above | 2 | 2.2% |
| Not stated | 9 | 10.1% |
| **Assessment at end of the study follow-up**<br>**(for final outcome) <sup>b</sup>** |  |  |
| Clinical assessment | 14 | 15.7% |
| Clinical and parasitological <sup>b</sup> | 65 | 73.0% |
| Clinical and/or parasitological <sup>b</sup> | 2 | 2.2% |
| Clinical and parasitological and molecular | 1 | 1.1% |
| Clinical and parasitological/serological | 1 | 1.1% |
| Not stated | 6 | 6.7% |
| **Time point of end of the study assessment** |  |  |
| < 6 months | 3 | 3.4% |
| 6 months | 75 | 84.3% |
| >6 to <12 months | 2 | 2.2% |
| 12 months | 8 | 9.0% |
| >12 months | 1 | 1.1% |
| **Definition of relapse** |  |  |
| Based on clinical suspicion | 4 | 4.5% |
| Clinical suspicion and/or parasitological demonstration | 2 | 2.2% |
| Parasitological demonstration upon clinical suspicion | 64 | 71.9% |
| Parasitological or serological | 1 | 1.1% |
| Molecular and/or Parasitological | 3 | 3.4% |
| Unclear | 15 | 16.9% |

<sup>a</sup> Achievement of clinical and parasitological resolution at the end of the treatment was usually defined as “initial cure”. Equivalent terminology adopted were: apparent cure, clinical and parasitological cure, apparent response or cure.
<sup>b</sup> Achievement of clinical and parasitological cure at the end of the study follow-up was usually defined as “definitive cure”. Equivalent terminology adopted were: full cure, final cure, definite cure, ultimate cure, complete cure or cure.

Tissue aspiration at TOC was done using spleen in 27 (30.3%), bone marrow in 5 (5.6%), lymph node in 2 (2.2%), a combination of bone marrow/spleen/lymph was used in 25 (28.1%), aspirate was carried out but the sample used was unclear in 16 (18.0%) studies, information regarding aspiration was not reported in 12 (13.5%), and aspiration was not done in 1 (1.1%) study (**Figure 3**).

**Figure 3:** Tissue aspiration used for confirmation of diseases and for outcome assessments Legend: See Supplemental file 2 for details for each of the studies included separately.

##### Duration of post treatment follow-up

Post-treatment follow-up duration was < 6 months in 3 (3.3%) trials, 6 months in 75 (84.3%), 7 months in 1 (1.1%), 9 months in 1 (1.1%), 12 months in 8 (9.0%) trials, and 2.8 years in 1 (1.1%) (**Table 4**).

Ability to complete the study follow-up was a criterion for inclusion in 21 (23.6%) studies, financial reimbursement of travel costs including food and logistics was provided in 6 (6.7%), active tracing of the patients either by visiting their home or by sending reminder by post/messenger was implemented in 11 (12.4%) studies, and there were no further details presented in the remaining 51 (57.3%) studies (**Supplemental file S2**).

##### Assessment of relapse and tissue aspiration

Relapse was defined solely based on clinical suspicion in 4 (4.5%) trials, parasitological demonstration upon clinical suspicion in 64 (71.9%) trials, based on molecular method (alone or in combination with parasitological method) in 3 (3.4%) studies, using clinical and/or parasitological assessments in 2 (2.2%), using parasitological or serological assessment in 1 (1.1%) study, and the definition of relapse was unclear in 15 (16.9%) studies.

Tissue aspiration used for confirmation of relapse included: splenic aspiration in 28 (31.5%) studies, bone marrow in 7 (7.9%) studies, bone marrow and/or spleen/lymph in 14 (15.7%) studies, peripheral blood in 1 (1.1%) study, tissue aspiration was done but the sample used was unclear in 30 (33.7%) studies, aspiration was not done in 3 (3.4%) studies and aspiration status was unclear in 6 (6.7%) studies (**Figure 3**).

#### Statistical considerations

##### Sample size calculation

Sample size calculation was carried out 34 (38.2%) studies, not carried out in 10 (11.2%) and there was no information in the remaining 45 (50.6%) studies (**Table 5**).

**Table 5:** Statistical considerations.

| Sample size estimation | Number of studies<br>(n=89 studies) | % |
| --- | --- | --- |
| **Objective of sample size estimation** |  |  |
| To detect difference in cure rate<br>(Difference: 8%-40%) | 13 | 14.6% |
| Demonstration of non-inferiority<br>(5%-20% non-inferiority margin) | 10 | 11.2% |
| To estimate cure rate with precision<br>(or 95% CI) | 4 | 4.5% |
| To detect desired efficacy/safety | 4 | 4.5% |
| To detect pharmacokinetics exposure | 1 | 1.1% |
| Unclear objective | 2 | 2.2% |
| Sample size calculation not carried out | 10 | 11.2% |
| No information presented regarding sample size estimation | 45 | 50.6% |
| **Adjustment of lost-to-follow up (LFU)**<br>**in sample size calculation** |  |  |
| Sample-size adjusted for possible LFU<br>(Adjustment: 5%-20%) | 11 | 12.4% |
| Not adjusted<br>(assumed no LFU) | 3 | 3.4% |
| Sample size calculation carried out but no information on LFU<br>adjustment | 20 | 22.5% |
| Sample size calculation not carried out | 10 | 11.2% |
| No information on sample size estimation | 45 | 50.6% |
| **Analysis approach** |  |  |
| Only PP explicitly reported | 2 | 2.2% |
| Only ITT explicitly reported | 14 | 15.7% |
| Both ITT and PP analysis reported | 27 | 30.3% |
| No explicit distinction/description of ITT or PP analysis | 46 | 51.7% |
| **Uncertainty interval (eg: 95% CI) presented along with point estimates of cured proportion?** |  |  |
| Point estimate presented with 95% CI | 50 | 56.2% |
| Only point estimate presented | 39 | 43.8% |
| **Was survival analysis used?** |  |  |
| Kaplan-Meier approach to estimate relapse | 3 | 3.4% |
| Kaplan-Meier approach to fever clearance | 2 | 2.2% |
| Not used | 84 | 94.4% |

ITT= Intention to treat; PP = Per protocol analysis; LFU = lost to follow-up

Of the 34 studies that reported sample size estimation, 2 studies reported sample size estimation based on safety endpoint, 1 based on pharmacokinetics endpoint, 1 based on a combination of safety and efficacy endpoint, 29 based on efficacy (or cure rate) endpoint, the endpoint was unclear in 1 study. In studies that presented the sample size estimation, 13 studies aimed to detect a mean difference in cured proportion (effect size: 8% to 40%), 10 studies aimed to demonstrate non-inferiority (margin adopted: 5% to 20%) and further details are presented in **Table 5**. Sample size adjustment for potential lost-to-follow up was carried out in 11 studies (adjustment range: 5% to 20%) and no adjustment was carried out in 3 studies (**Table 5**).

##### Analysis approach: ITT or PP

Intention-to-treat analysis was undertaken in 41 studies and this was not clear in the remaining 48 studies. Per-protocol analysis was undertaken in 29 studies and this was not clear in the remaining 60 studies. Both ITT and PP analysis was undertaken in 27 studies. In 46 studies, there were no explicit description of the ITT or PP analysis (**Table 5**).

##### Estimation of drug efficacy

Cured proportion at the end of the study follow-up was presented in all 89 studies included. The point estimate of cured proportion was presented along with an interval estimates (95% confidence interval for the cured proportion or for the mean difference in proportion between two groups) in 50 (56.2%) studies and only point estimates were presented in the remaining 39 studies. Survival analysis was used in 5 (5.6%); 3 studies used Kaplan-Meier approach for estimating incidence of relapse while 2 studies used for estimating fever clearance (**Table 5**).

## Discussion

Our review has characterised the variations in design, conduct, analysis, and reporting of prospective therapeutic efficacy studies conducted in VL for the last 20 years. The patient flow diagram was presented in only 46 of the 89 studies included in this review; a third of those who were screened for eligibility were excluded in studies that clearly presented the details. The most commonly reported reason for exclusion was negative parasitaemia upon further parasitological/serological examination among third of the excluded patients suggesting a need for a more sensitive case definition for identification of VL patients. For patient enrolment, those who met the case definitions underwent tissue examination with splenic aspirates as the most commonly used tissue sample; approximately a third of the studies used splenic aspirates for patient enrolment and further two-thirds used splenic aspiration in combination with bone marrow. In patients with low-grade parasitaemia, non-palpable spleen can lead to difficulty in splenic aspiration and thus studies generally rely on bone marrow aspirates that have lower sensitivity. A reliable confirmatory diagnosis of VL can be challenging and this urgently necessitates the development of a highly sensitive non-invasive sampling approach such as the adoption of molecular tools ^12^. A recent evaluation found that recombinase polymerase amplification had a high concordance with polymerase chain reaction based on peripheral blood sample^13^; such tool can serve as an alternative approach for disease confirmation and monitoring parasite load.

There was a notable variability in the inclusion and exclusion criteria adopted including the ranges of haemoglobin, platelets and other liver enzymes tests, and co-infection with major comorbidities such as HIV, TB, hepatic, cardiac and renal complications. Overall, these criteria will generally exclude the patients with critical ill patients or those with severe form of VL. Similarly, pregnant and lactating women were excluded in 60% of the studies. Overall, this suggests the need for innovative trial designs to provide further information regarding the drug effectiveness in population the patient population excluded from the standard therapeutic efficacy studies. For example, responsible inclusion of these patients in a pragmatic trial ^14^ or the use of registries or observational databases might facilitate further assessment of drug effectiveness among these excluded population.

There were several elements of the studies that were poorly reported. For example, 40% of the randomised studies did not report the approaches used for sequence generation and 45% did not report on the allocation concealment. Age-range of the patients included was not reported in a fifth of the studies included in this review, and there was no statement regarding requirement of informed consent in 11 of the 89 studies. Information regarding the quality control of laboratory procedures adopted in the studies were not reported in the majority of the studies, and the primary endpoint of the study was not explicitly stated in 44% of the included studies. And when stated and reported, several different terminologies were used for referring to the same treatment endpoints. For example, initial cure was most commonly defined as a composite of initial parasitological and clinical cure at the end of the therapy period in majority of the studies; alternative terminology adopted included initial apparent cure, apparent response or cure (supplemental file 2). Definitive cure assessed at the end of the study follow-up required demonstration of clinical and parasitological cure with absence of relapse after achieving initial cure; alternative terminologies used included ultimate cure, definite cure, final cure, and full cure (**supplemental file 2**). Similarly, in 46 (43.8%) studies, only point estimates of the cured proportion were presented without presenting the associated uncertainty estimates. Only a third of the studies clearly reported the efficacy estimates from both ITT and PP analysis while there was no specific distinction between these two approaches in over half of the studies (Table 5). These findings are consistent with a previous review that identified that VL relapses were not adequately defined ^15^ and suggests an urgent need for a harmonisation of terminologies and reporting practices.

There were also variations in the time-point when the drug efficacy was monitored. Majority of the studies assessed the test of cure within 30 days of treatment completion and assessed definitive cure at 6-months. In particular, the assessment of definitive cure requires a careful evaluation as this requires absence of relapse. Field observations have reported that a substantial proportion of patients develop relapse after 6-months ^16,17^ suggesting that a 6-months follow-up duration may be overestimating efficacy^18^. Asymptomatic relapse as observed in a trial from India ^19^, can pose further challenges as the current algorithm for detection of relapse is conditional upon a patient showing clinical signs and symptoms. Asymptomatic relapses or those with low grade parasite load with non-palpable spleen can easily misclassified as definitive cure instead of relapse. This can have important ramifications for providing an infective parasite pool for fuelling further transmission of the disease. It is thus important to develop an accurate diagnostic algorithm for identifying relapses; such algorithm can be a complementary tool to aid-in the diagnosis in settings where it might be difficult to carry out aspiration or if following-up patients might not be possible.

Development of a clear definition of suspected VL, differentiating severe VL from uncomplicated/moderate disease, defining relapse and suspected case of relapse, and a standardised protocol on design and conduct of VL studies would help in harmonisation of clinical practices, as has been proposed for cutaneous leishmaniasis ^5,6^. Although country specific treatment guidelines exist for treatment of patients under field conditions, to date, there is a clear lack of such protocol for VL. For example, recently a checklist has been developed for reporting of malaria therapeutic efficacy studies in collaboration between researchers and academic editors of the journals ^20^. Lessons learnt from such initiative can be applied in the context of VL will help in reducing the reporting heterogeneity in VL studies. The recent development of CIDISC standards for capturing data in clinical studies through the IDDO-DND*i* collaboration and the development of standardised case report forms are an important steps towards harmonisation of standards ^21^. Such harmonisation is important to maximising the information from limited trials as conducting therapeutic studies for novel drugs remains increasingly challenging in the Indian sub-continent due to the falling burden of the disease ^22^ and in East Africa due to political instability ^23^.

Some of the challenges identified in this review and potential solutions are summarised in **Box 1**. A reporting checklist has been developed based on the completeness of reporting of different aspects of trial design, conduct, analysis and reporting identified in this review and is presented in **Box 2**.

## Conclusions

This review highlights substantial methodological variations in definitions adopted for patient screening, disease diagnosis and therapeutic outcomes suggesting a need for a harmonised protocol for design and conduct of VL clinical studies.

## Declarations

### Authors’ contributions

Conceptualization: PD, SSP, PJG, KS

Data Curation: PD, SSP

Formal Analysis: PD, SSP, MC, PJG, KS

Funding Acquisition: PJG

Investigation: PD, SSP, PJG, KS

Methodology: PD, SSP, PJG, KS

Project Administration: MB, CN, SSP, PJG

Resources: PJG Software: PD

Supervision: PJG, KS

Validation: PD, SSP, PJG, KS

Visualization: PD

Writing – Original Draft Preparation : PD, SSP, FA, PJG, and KS

Writing – Review & Editing : All

### Availability of data and material

The database(s) supporting the conclusions of this article are available within the tables and figures presented within the manuscript along with the supplemental files (S1 and S2).

### Ethics approval and consent to participate

Not applicable

### Consent for publication

Not applicable

## Box 1.

### Key methodological challenges and potential solutions

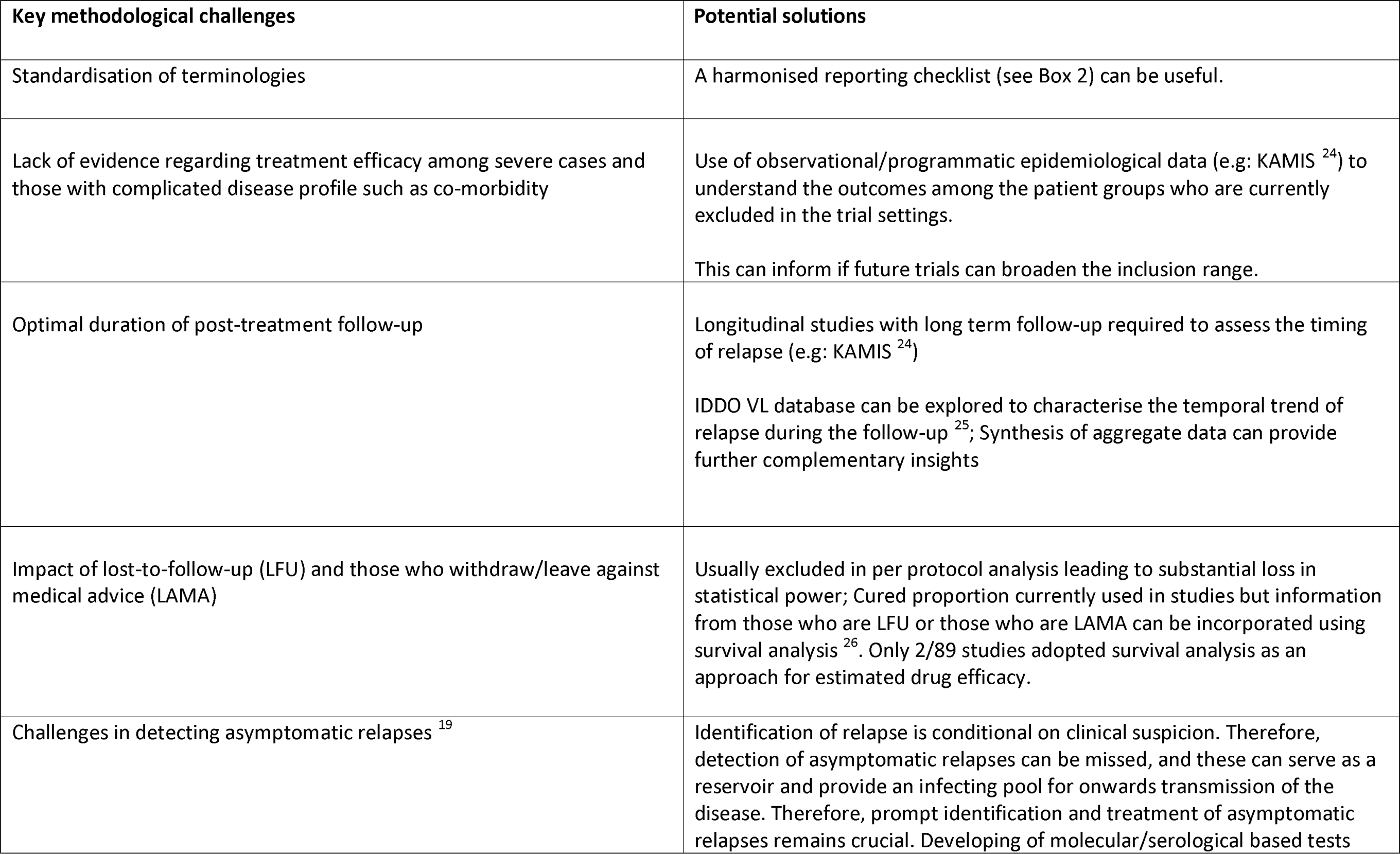
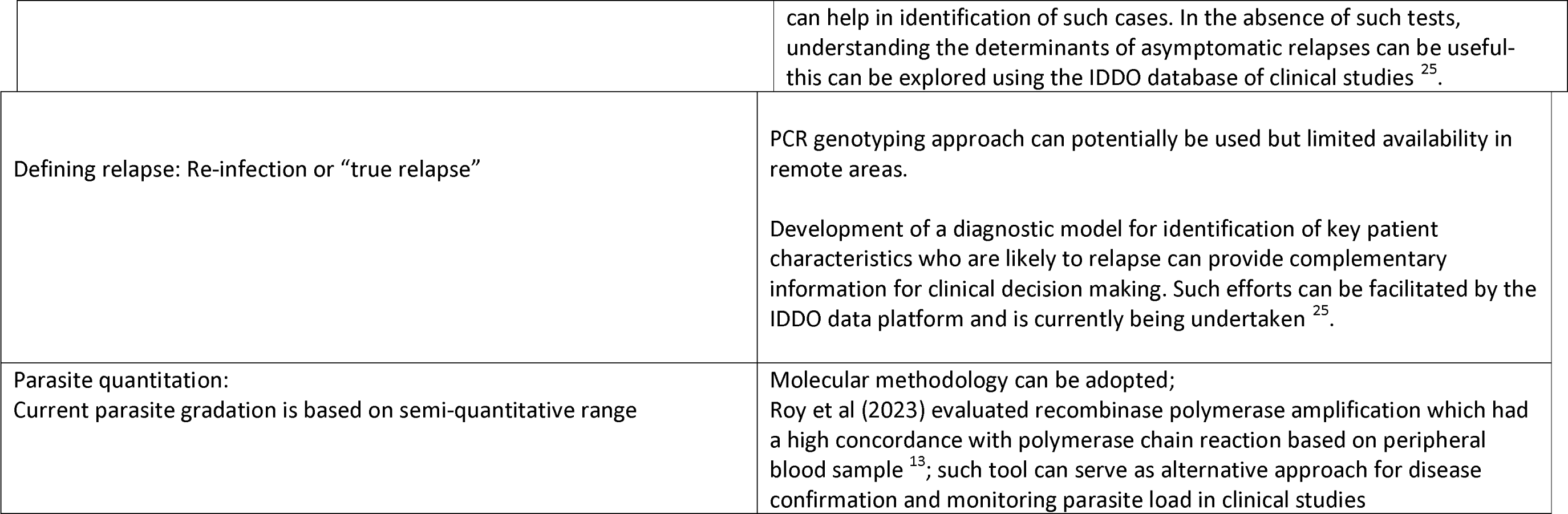

## Box 2.

### Suggested reporting checklist for therapeutic efficacy studies in visceral leishmaniasis

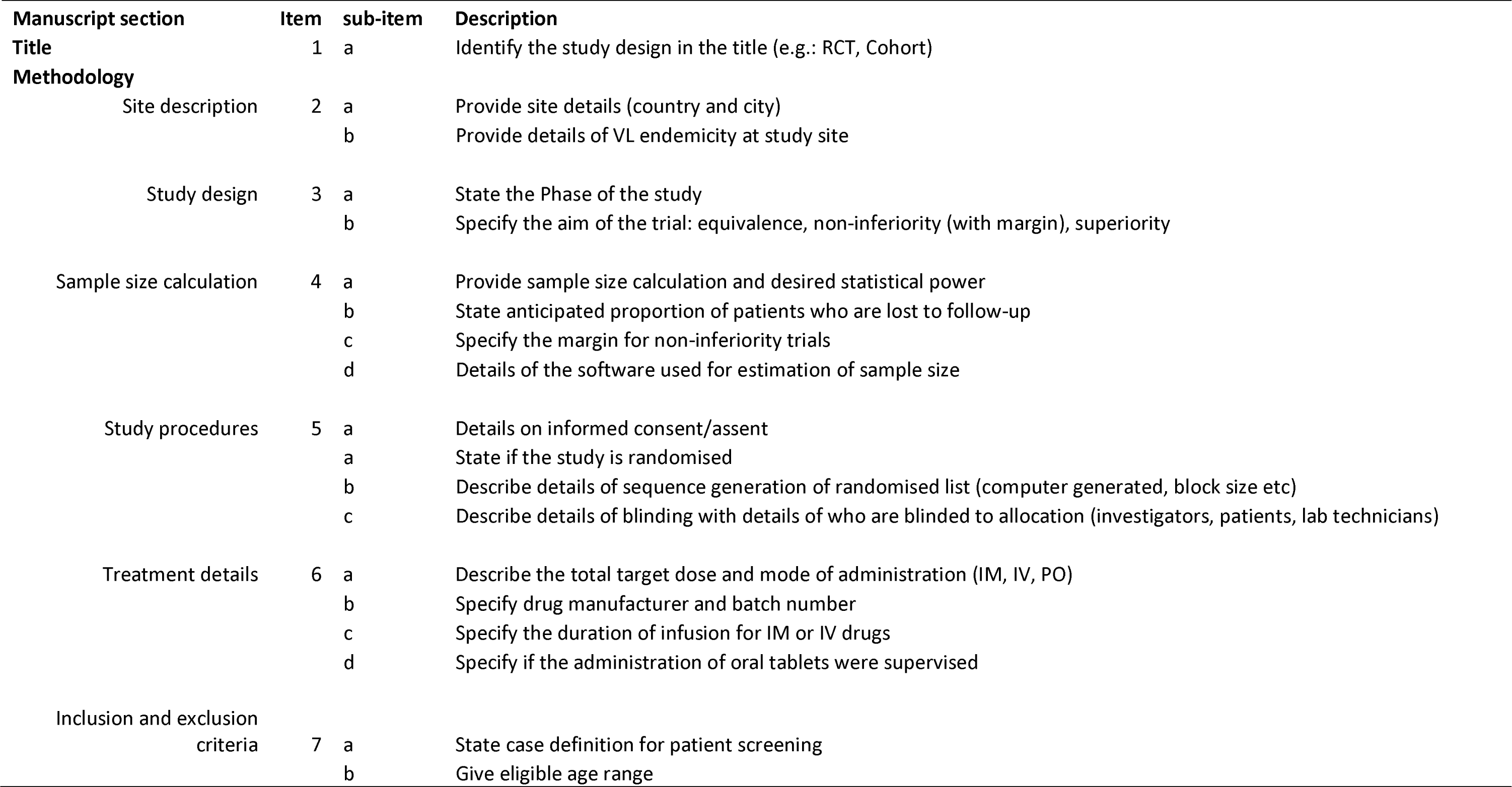
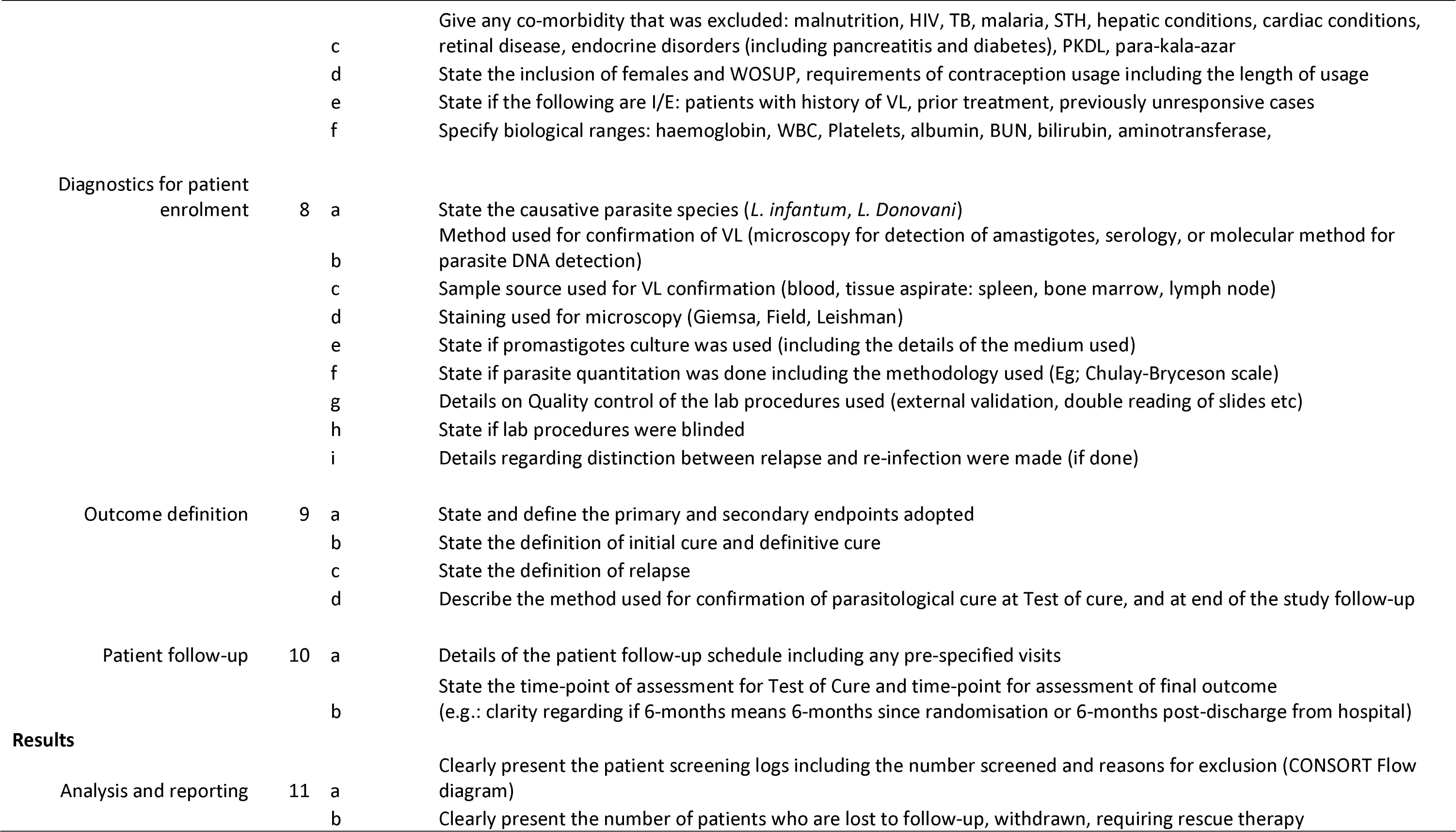
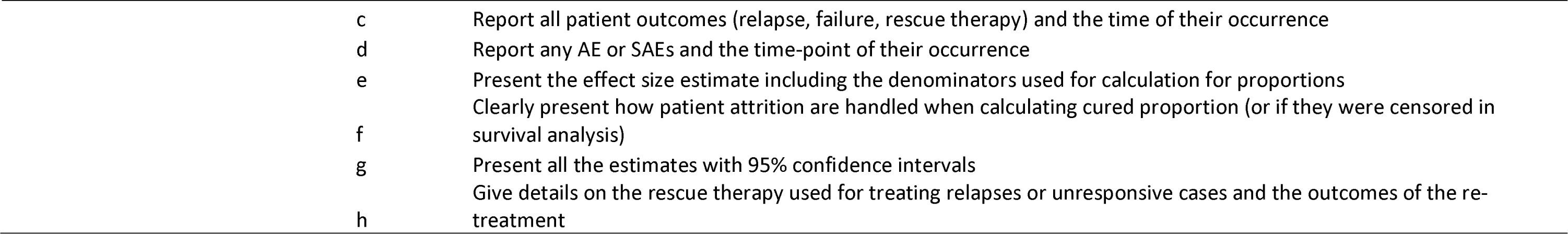

