## Supplemental file 1 for "Design, conduct, analysis, and reporting of therapeutic efficacy studies in Visceral Leishmaniasis: A systematic review of published reports, 2000-2021"

**Supplemental file S1: Study details**

**Table 1: List of studies included in the review**

| **Author-year** | **PubMed** | **Country** | **Randomisation** | **Follow-up**  **(days)** | **Enrolled** | **Treatment**  **blinding** | **Age range** | **Patients with HIV co-infections** | **Pregnant and lactating** |
| --- | --- | --- | --- | --- | --- | --- | --- | --- | --- |
| Gaeta-2000 | 11200380 | Italy | Single Group | 180 | 12 | Unclear | All ages | Excluded | Not clear |
| Sundar-2000 | 11049800 | India | Randomised | 180 | 54 | Open | All ages | Excluded | Excluded |
| Thakur-2000 | 11127250 | India | Randomised | 180 | 150 | Open | All ages | Not clear | Not clear |
| Sundar-2000 | 11049798 | India | Single Group | 180 | 320 | Unclear | All ages | Not clear | Not clear |
| Sundar-2000 | 10897369 | India | Single Group | 180 | 70 | Unclear | All ages | Excluded | Excluded |
| Thakur-2000 | 11127251 | India | Randomised | 180 | 120 | Open | All ages | Not clear | Excluded |
| Veeken-2000 | 10886792 | Sudan | Non-randomised | 180 | 516 | Open | All ages | Not clear | Not clear |
| Villanueva-2000 | 11117648 | Not stated | Non-randomised | 1020 | 32 | Unclear | Not clear | Included | Not clear |
| Thakur-2001a | 11137652 | India | Randomised | 180 | 34 | Unclear | All ages | Excluded | Excluded |
| Moore-2001 | 11417033 | Kenya | Non-randomised | 180 | 102 | Open | All ages | Not clear | Not clear |
| Ritmeijer-2001 | 11816442 | Ethiopia | Non-randomised | 180 | 199 | Open | All ages | Included | Not clear |
| Thakur-2001b | 11280166 | India | Single Group | 180 | 309 | Unclear | All ages | Not clear | Not clear |
| Dietze-2001 | 11791957 | Brazil | Non-randomised | 360 | 22 | Open | All ages | Excluded | Excluded |
| Sundar-2001 | 11520836 | India | Randomised | 180 | 91 | Open | Not clear | Excluded | Excluded |
| Haidar-2001 | 11426243 | Yemen | Single Group | 20 | 32 | Unclear | Less than 15y | Not clear | Not applicable |
| Das-2001 | 11584934 | India | Randomised | 180 | 158 | Unclear | All ages | Not clear | Not clear |
| Sundar-2002a | 12456849 | India | Randomised | 180 | 398 | Open | All ages | Excluded | Excluded |
| Sundar-2002b | 12135284 | India | Randomised | 180 | 84 | Blinded | All ages | Excluded | Excluded |
| Laguna-2003 | 12888588 | Spain | Randomised | 180 | 57 | Open | Adults | Included | Excluded |
| Sundar-2003 | 12955641 | India | Single Group | 180 | 203 | Open | All ages | Excluded | Excluded |
| Figueras Nadal-2003 | 14636517 | Spain | Single Group | 180 | 32 | Open | Less than 15y | Excluded | Not clear |
| Sundar-2003 | 12792385 | India | Non-randomised | 180 | 39 | Open | Less than 15y | Excluded | Not applicable |
| Syriopoulou-2003 | 12594635 | Greece | Non-randomised | 180 | 123 | Open | Less than 15y | Excluded | Not applicable |
| Rijal-2003 | 15228258 | Nepal | Single Group | 180 | 120 | Open | All ages | Excluded | Not clear |
| Thakur-2004 | 15035723 | India | Not Specified | 180 | 120 | Unclear | All ages | Excluded | Not clear |
| Sundar-2004 | 14727208 | India | Randomised | 180 | 153 | Open | All ages | Excluded | Excluded |
| Thakur-2004b | 15489554 | India | Not Specified | 180 | 282 | Unclear | All ages | Excluded | Not clear |
| Bhattacharya-2004 | 14699453 | India | Single Group | 180 | 80 | Open | Less than 15y | Excluded | Excluded |
| Wasunna-2005 | 16282296 | Kenya | Non-randomised | 180 | 97 | Open | All ages | Excluded | Excluded |
| Jha2005 | 16354802 | India | Randomised | 180 | 120 | Open | All ages | Excluded | Excluded |
| Das2005 | 16130613 | India | Single Group | 180 | 182 | Unclear | All ages | Not clear | Not clear |
| Ritmeijer-2006 | 16804852 | Ethiopia | Randomised | 180 | 580 | Open | All ages | Included | Not applicable |
| Sundar-2006 | 16447104 | India | Randomised | 180 | 405 | Open | All ages | Excluded | Excluded |
| Singh-2006 | 17202605 | India | Single Group | 180 | 64 | Unclear | Less than 15y | Excluded | Not applicable |
| Singh-2006b | 17202633 | India | Randomised | 180 | 125 | Unclear | Less than 15y | Excluded | Not applicable |
| Mueller-2006 | 16730363 | Sudan | Single Group | 30 | 64 | Unclear | All ages | Included | Excluded |
| Sundar-2007 | 17682988 | India | Randomised | 180 | 1485 | Open | All ages | Excluded | Excluded |
| Sundar-2007b | 17582067 | India | Randomised | 180 | 667 | Open | All ages | Excluded | Excluded |
| Bhattacharya-2007 | 17624846 | India | Single Group | 180 | 1132 | Open | All ages | Excluded | Excluded |
| Sundar-2008 | 18781879 | India | Randomised | 270 | 226 | Open | All ages | Excluded | Excluded |
| Sundar-2008b | 18664241 | India | Non-randomised | 180 | 45 | Open | All ages | Excluded | Excluded |
| Thakur-2008 | 18765878 | India | Randomised | 180 | 140 | Blinded | All ages | Not clear | Not clear |
| Mueller-2008 | 18186974 | Uganda | Non-randomised | 180 | 371 | Unclear | All ages | Not clear | Not clear |
| Sundar-2009 | 19407109 | India | Non-randomised | 180 | 60 | Open | Adults | Excluded | Excluded |
| Das-2009 | 19436614 | India | Randomised | 180 | 82 | Open | All ages | Excluded | Excluded |
| Sundar-2009b | 19663597 | India | Randomised | 180 | 329 | Open | All ages | Excluded | Excluded |
| Adam-2009 | 19766208 | Sudan | Single Group | 365 | 42 | Unclear | All ages | Excluded | Excluded |
| Shahian-2009 | 19478699 | Iran | Single Group | 180 | 20 | Unclear | Less than 15y | Excluded | Not applicable |
| Hailu-2010 | 21049059 | Sudan, Kenya, Ethiopia | Randomised | 180 | 270 | Open | All ages | Not clear | Excluded |
| Thakur-2010 | 21036834 | India | Randomised | 360 | 230 | Open | All ages | Excluded | Not clear |
| Musa-2010 | 21049063 | Sudan | Randomised | 180 | 42 | Open | All ages | Excluded | Excluded |
| Sundar-2010 | 20147716 | India | Randomised | 180 | 412 | Open | All ages | Excluded | Not clear |
| Mondal-2010 | 20668544 | India | Randomised | 180 | 25 | Open | All ages | Excluded | Excluded |
| Rijal-2010 | 19726065 | Nepal | Single Group | 180 | 198 | Unclear | All ages | Not clear | Not clear |
| Singh-2010 | 20065047 | India | Randomised | 180 | 605 | Unclear | Less than 15y | Excluded | Not applicable |
| Sinha-2010 | 20682882 | India | Single Group | 180 | 251 | Unclear | All ages | Excluded | Excluded |
| Sundar-2011 | 21255828 | India | Randomised | 180 | 634 | Open | All ages | Excluded | Excluded |
| Sundar-2011b | 21633025 | India | Randomised | 180 | 61 | Open | All ages | Excluded | Excluded |
| Sundar-2011c | 21129762 | India | Single Group | 180 | 135 | Unclear | All ages | Excluded | Excluded |
| Rahman-2011 | 21734127 | Bangladesh | Single Group | 180 | 977 | Open | All ages | Excluded | Excluded |
| Sinha-2011 | 22174722 | India | Single Group | 180 | 494 | Open | All ages | Excluded | Excluded |
| Sudarshan-2011 | 21609983 | India | Randomised | 46 | 46 | Unclear | All ages | Excluded | Not clear |
| Sundar-2012 | 22573856 | India | Single Group | 180 | 567 | Open | All ages | Excluded | Excluded |
| Musa-2012 | 22724029 | Sudan, Ethiopia, Kenya, Uganda | Randomised | 180 | 972 | Open | All ages | Included | Excluded |
| Patra-2012 | 23087513 | India | Single Group | 180 | 71 | Unclear | All ages | Excluded | Excluded |
| Rijal-2013 | 23425958 | Nepal | Single Group | 360 | 120 | Unclear | All ages | Included | Excluded |
| Khalil-2014 | 24454970 | Ethiopia, Sudan | Randomised | 180 | 124 | Open | Not clear | Excluded | Excluded |
| Ostyn-2014 | 24941345 | India, Nepal | Single Group | 360 | 1016 | Unclear | All ages | Excluded | Not clear |
| Cota-2014 | 24743472 | Brazil | Non-randomised | 180 | 90 | Unclear | All ages | Included | Not clear |
| Sundar-2014 | 25233346 | India | Randomised | 180 | 500 | Open | All ages | Excluded | Not clear |
| Mondal-2014 | 25104636 | Bangladesh | Single Group | 180 | 300 | Unclear | All ages | Not clear | Excluded |
| Jamil-2015 | 26496648 | Bangladesh | Single Group | 180 | 120 | Open | All ages | Excluded | Excluded |
| Sundar-2015 | 25510715 | India | Non-randomised | 180 | 30 | Open | All ages | Excluded | Excluded |
| Goswami-2016 | 26526926 | India | Randomised | 180 | 120 | Open | All ages | Excluded | Excluded |
| Wasunna-2016 | 27627654 | Kenya, Sudan | Randomised | 210 | 183 | Open | All ages | Excluded | Not applicable |
| Pandey-2016 | 27645786 | India | Single Group | 270 | 646 | Open | All ages | Excluded | Excluded |
| Borges-2017 | 28327804 | Brazil | Randomised | 180 | 101 | Open | Less than 15y | Not clear | Not applicable |
| Rahman-2017 | 28558062 | Bangladesh | Randomised | 180 | 602 | Open | All ages | Excluded | Excluded |
| Romero-2017 | 28662034 | Brazil | Randomised | 180 | 378 | Open | All ages | Excluded | Excluded |
| Alborzi-2017 | 27879460 | Iran | Randomised | 180 | 75 | Open | All ages | Not clear | Not clear |
| Pandey-2017 | 29016288 | India | Single Group | 180 | 100 | Open | Not clear | Excluded | Not clear |
| Kimutai-2017 | PMC5315726 | Sudan, Ethiopia, Kenya, Uganda | Single Group | 180 | 3126 | Open | All ages | Included | Included |
| Mbui-2018 | 30188978 | Kenya, Uganda | Single Group | 210 | 30 | Open | Less than 15y | Excluded | Excluded |
| Goyal-2018 | 30346949 | India | Non-randomised | 180 | 1761 | Open | All ages | Excluded | Included |
| Sundar-2019 | 31436156 | India | Single Group | 360 | 1143 | Unclear | All ages | Excluded | Not Clear |
| Diro-2019 | PMC6336227 | Ethiopia | Randomised | 58 | 58 | Open | Adults | Included | Excluded |
| Sinha-2019 | Not indexed | India | Single Group | 208 | 160 | Open | Less than 15y | Excluded | Excluded |
| Goswami-2020 | 32394874 | India | Randomised | 1825 | 154 | Open | All ages | Excluded | Excluded |
| Ekram-2021 | 34789971 | Bangladesh | Single Group | 180 | 31 | Open | All ages | Excluded | Excluded |

**Table 2: Details of randomisation methods adopted in the studies included**

| **Author-year** | **Randomisation details** | **Sequence generation** | **Allocation Concealment** |
| --- | --- | --- | --- |
| Ritmeijer-2006 | The patient was randomized to receive miltefosine or SSG according to a computer-generated number list. The allocation ratio was 1:1. The study was unblinded; miltefosine is oral medication and SSG is injection medication. | Computerised | Unclear |
| Thakur-2001a | Matched for age and sex, the patients were randomly allocated into two treatment groups. | Unclear | Unclear |
| Laguna-2003 | A randomization list was prepared using the SAS program, which stratified patients into two groups, depending on the CD4 cell count at inclusion: above or below 200 cells/mm3. If this information was missing at the time of randomization, it was considered equivalent to the lympho- cyte count: above or below 1000 cells/mm3. The randomization process was blinded and centralized. | Computerised | Unclear |
| Sundar-2008b | An independent statistician generated a randomization schedule by use of a computer-based procedure; assumptions were a maximum number of 60 patients enrolled per arm (240 total patients) and 15 randomization blocks with a size of 15 patients each. Sealed randomization envelopes were prepared, and treatment was begun within 72 h after diagnosis by splenic aspirate. | Computerised | Sealed envelope |
| Sundar-2002b | Patients were centrally registered and randomly assigned at each site to miltefosine or amphotericin therapy in a 3:1 ratio with the use of permuted blocks of four patients each. | Unclear | Unclear |
| Sundar-2000c | After completing initial diagnostic and baseline laboratory testing, 54 patients were randomized, by means of a computer- generated, sealed-envelope method, to receive one 50-mg capsule of miltefosine twice daily with meals for either 14 (group A), 21 (group B), or 28 days (group C). | Computerised | Sealed envelope |
| Das-2009 | Treatment allocation was done by the biostatistician of the institute, who performed the allocation sequence using random number tables and accordingly assigned the test and control group. A total of 82 SSG unresponsive and parasitologically confirmed VL cases were divided randomly into two groups before the initiation of the treatment. | Random number table | Unclear |
| Thakur-2000a | Unclear | Unclear | Unclear |
| Jha-2005 | Patients were randomized contemporaneously to receive sitamaquine daily for 28 days at one of four doses. The randomization schedule provided for an equal number of subjects in all four cohorts (N⳱ 30). However, to minimize the number of patients exposed to higher sitamaquine doses, the randomization schedule was not followed for the final block of 8 subjects (Subjects 113 to 120). These 8 subjects were entered into Cohorts 1 and 2. | Unclear | Unclear |
| Sundar-2006 | An independent statistician prepared randomization envelope by use of a computer-generated random-number generator. The sealed envelopes were then distributed to the enrolled subjects, to randomly assign them to 1 of the 3 fol- lowing ABCD total-dose groups: 7.5 mg/kg (group A), 10 mg/ kg (group B), and 15 mg/kg (group C). | Computerised | Sealed envelope |
| Sundar-2004 | An independent statistician prepared randomization envelope using a computer- based random number generator. Enrolled subjects were randomly assigned by sealed envelope to receive 3 treatments | Computerised | Sealed envelope |
| Sundar-2007b | For the purposes of this study, we considered 15 alternate-day infusions of 1 or 0.75 mg/kg (groups A and B) as conventional therapy [2, 3, 5, 8]. To compare responses to these same doses given once daily (groups C and D), we used a 1:2 ratio for random assignment to treatment arms and aimed to enrol 250 subjects each in groups A and B and 500 subjects each in groups C and D. An independent statistician prepared sealed randomization envelopes using a computer-based random number generator. | Computerised | Sealed envelope |
| Thakur-2008 | Of the 181 patients screened, only 140 met the inclusion and exclusion criteria and gave written consent. These were divided randomly in two groups with 70 patients, each matched for age and sex; 55 men and 15 women were included in each group (Fig. 2). As there was some difficulties in culturing grade-1 amastigotes, patients with grade-1 amastigotes were excluded from both the groups to maintain parity. | Unclear | Unclear |
| Sundar-2007a | Enrolled patients were randomly assigned to treatment with paromomycin or amphotericin in a 3:1 ratio in permuted blocks of four. A fraction of the patients in the paromomycin group were also randomly assigned to a sub study in which pharmacokinetic sampling was performed. | Unclear | Unclear |
| Sundar-2002a | Patients were randomized into preassigned treatment groups by the sealed-envelope technique, and liposomal amphotericin B was administered by an independent coinvestigator who broke the seal of the envelope and prepared infusions | Unclear | Sealed envelope |
| Sundar-2009b | Patients were allocated to a randomization arm from a random table generated for this purpose. | Unclear (random number tables) | Unclear |
| Sundar-2001 | participants were randomised by sealed envelope to receive 5 mg/kg of liposomal amphotericin (AmBisome, NeXstar, Paris) as a single infusion or as once daily infusions of 1 mg/kg for five consecutive days. An independent statistician prepared the randomisation envelopes using a computer-based random number generator. The study staff opened consecutively numbered envelopes containing the treatment assignment after eligible patients fulfilled the entry criteria. | Computerised | Sealed envelope |
| Thakur-2000b | Patient eligibility was evaluated before randomisation to treatment with a computer-generated randomisation list. | Computerised | Unclear |
| Singh-2006b | Patients were randomized into four groups by slips kept separately in two small boxes for both newly diagnosed patients and those who had received 30 days course of sodium antimony gluconate (SAG): | Unclear | Boxed |
| Das-2001 | The patients were randomly allocated to two regimen groups, combination regimen of pentamidine (half dose) and allopurinol, and single regimen of pentamidine alone. | Unclear | Unclear |
| Sundar-2011b | A computer-generated, randomisation code was generated by the trial statistician by use of SAS 9.1 (SAS Institute, Cary, NC, USA). To ensure maximum balance of the numbers in each group at any time and to minimise bias, block sizes of 16 were generated for treatment allocation of patients to one of the four treatment groups, with equal allocation ratio and independently for each site. Individual, opaque, sealed, and sequentially numbered envelopes were provided to each trial site, one envelope per patient, to indicate the allocation of individual patients to treatment. | Computerised | Sealed envelope |
| Sundar-2011c | Patients were randomized into blocks of 12 in the ratio 1:1:1:1:2 to one of four sitamaquine cohorts or AmB ( Figure 1 ). Treatment was allocated by using the GlaxoSmithKline Registration and Medication Ordering System (RAMOS). | Computerised | Unclear |
| Hailu-2010 | Allocation to treatment was by means of sequentially numbered, sealed envelopes, generated from a computerized randomization list. Each centre received a box of uniquely numbered sealed envelopes from the LEAP Trial Coordination Centre in Nairobi, where centralized randomization and envelope preparation were carried out in blocks of 15 to maintain randomization balance within centres. | Computerised | Sealed envelope |
| Thakur-2010 | This study was conducted as an open-label, randomized trial of 230 patients at Balaji Utthan Sansthan, Patna. The study staff who treated the patients opened consecutively numbered envelopes containing the treatment assignment after eligible patients fulfilled the entry criteria. Clinicians who provided treatment were not blinded to the treatment given. | Unclear | Sealed envelope |
| Musa-2010 | Randomization was done using sequentially numbered sealed envelopes that were prepared according to a centrally generated randomization list. | Unclear | Sealed envelope |
| Khalil-2014 | Patients were randomized to receive either treatment using a computer-generated randomisation list, stratified by site. Individual treatment allocations were placed in sealed, opaque envelopes which were opened after a patient had been entered into the trial. | Computerised | Sealed envelope |
| Sundar-2010 | To compare responses to liposomal amphotericin B versus amphotericin B deoxycholate, we used a 3:1 ratio for random assignment to treatment, aiming to assign 300 patients to receive liposomal amphotericin B (liposomal-therapy group) and 100 to receive amphotericin B deoxycholate (conventional-therapy group). An independent statistician prepared sealed randomization envelopes, using a computer-based random-number generator. | Computerised | Sealed envelope |
| Musa-2012 | A computer-generated randomization list was produced with stratification by centre and block sizes of 15 until recruitment in the PM arm was completed, and block sizes of 10 thereafter. Allocation was concealed using opaque, sequentially numbered sealed envelopes. The randomization list and envelopes were prepared and stored securely at the LEAP Data Centre, based at the trial coordination centre in Nairobi. | Computerised | Sealed envelope |
| Mondal-2010 | An independent statistician prepared randomization envelops by the use of a computer-generated random number table. The sealed envelopes were then distributed to the enrolled subjects, to randomly assign them to one of the following total dose groups; 5 mg/kg single shot (n = 10), 7.5 mg/kg single shot (n = 10) and 5 mg/kg double shot (total 10 mg/kg) (n = 10) Fungisome. | Computerised | Sealed envelope |
| Singh-2010 | They were randomized into two treatment groups, Groups A and B by electronically generated random table | Computerised | Unclear |
| Sudarshan-2011 | Not provided | Unclear | Unclear |
| Goswami-2016 | Patients were randomized to receive either treatment using a computer-generated randomization list. Individual treatment allocations were placed in sealed, opaque envelopes, which were opened after a patient had been entered into the trial. It was not possible to blind patients or treating physicians because of the nature of the intervention. | Computerised | Sealed envelope |
| Sundar-2014 | The permuted block randomization, with block size of 4, and ratio of 3:1 in the two groups (ABLE and LAmB) were generated for each center. Eligible patients were sequentially allotted to unique subject ID and treatment (ABLE or LAmB) as per randomization schedule for that center. The screening and randomization log were maintained. | Unclear | Unclear |
| Diro-2019 | Subjects were allocated to treatment using random block sizes, stratified by site (Gondar & Abdurafi) and by patient type (whether the VL episode at screening was a primary or relapse case). The randomization list was prepared by the data management team. Site investigators were blinded to block sizes. Randomization codes were prepared in sealed, sequentially numbered, opaque envelopes and were under the control of the site investigator. | Unclear | Sealed envelope |
| Borges-2017 | Randomization procedure was performed in blocks of 20, using Graphpad Quickcalcs free software (GraphPad Software Inc., San Diego, CA). This procedure was under the responsibility of an independent researcher from Tropical Medicine Center at Universidade de Brasília, who was not directly involved in any other operational aspect of the study. The names of compared drugs were typed on a sheet and placed in dark envelopes, which were sealed, stamped, and signed by those responsible for the randomization. The envelopes were opened immediately after the consent of the participants. | Computerised | Sealed envelope |
| Wasunna-2016 | Subjects were randomly allocated using block randomization, stratified by site (Dooka, Kassab and Kimalel). Site investigators were blinded to block size and codes were concealed in sealed sequentially numbered, opaque envelopes under the control of the site investigator. | Unclear | Sealed envelope |
| Rahman-2017 | A computer-generated randomization code was used for patient treatment allocation. Individ- ual, opaque, sealed and sequentially numbered envelopes were provided to each study site (one envelope per patient), indicating the individual patient allocation to treatment. | Computerised | Sealed envelope |
| Romero-2017 | Computer-generated randomization into the four treatment arms was done using the software Quickcalcs-online calculators for scientists (www.graphpad.com/quickcalcs/randomize1.cfm). Blocks of28 treatment allocations were generated and placed in sealed, opaque envelopes that were sent to each clinical trial site, and only opened by trial clinicians or site investigators when a participant was included in the trial. After the AmphoB arm was withdrawn, ifan enrolled patient was allocated to this treatment arm, the subsequent envelope containing a new code was designated to the patient until they were allocated to any ofthe three remaining arms. This type ofapproach did not allow for blinding. | Computerised | Sealed envelope |
| Goswami-2020 | A computer-generated, randomization code was generated. Enrolled patients were randomly assigned to treatment with miltefosine or with the combination chemotherapy in a 1:1 ratio. Microscopists were masked to the treatment given. | Computerised | Unclear |
| Alborzi-2017 | The patients were randomized into three groups regardless of age, sex, and duration of the disease. | Unclear | Unclear |
