## Supplemental file 2 for "Design, conduct, analysis, and reporting of therapeutic efficacy studies in Visceral Leishmaniasis: A systematic review of published reports, 2000-2021"

**Supplemental file S2: Further tables**

**Table 1: Inclusion and exclusion criteria adopted in the studies**

| **Author year** | **Inclusion/Exclusion criteria** |
| --- | --- |
| Gaeta-2000 | HIV-negative individuals over 18 y were eligible if Leishmania was visible in, or cultured from, a bone marrow aspirate |
| Sundar-2000c | Inclusion and exclusion criteria and baseline laboratory tests (as well as all trial procedures) were identical to those used in our prior studies of miltefosine and are described in detail elsewhere. Patients were excluded from the study if they had another serious concurrent disease or infection or if they were <12 years of age or >65 years of age, were pregnant or breast-feeding, or were determined to be seropositive for HIV by means of ELISA testing (no screened subject was seropositive). Other exclusion criteria were a WBC count of <2000 cells/mm3, a haemoglobin level of <6 g/dL, a platelet count of <50,000 cells/mm3, levels of hepatic transaminases or total bilirubin >3 times the normal levels, a serum creatinine level of>2 mg/dL, and a prothrombin time >5 s above the control value. |
| Thakur-2000a | Refer to JHA et al. (1998) (IDDO TAG 69) for a complete description of materials and methods employed in this study.  From Jha-1998: Exclusion criteria were a known allergy to aminoglycosides, treatment in the previous 12 months with a drug with recognised or presumed anti-leishmanial action, serious concomitant diseases, pregnancy or lactation (women underwent a pregnancy test at initial assessment), failure to agree to return for all follow up evaluations, and being critically ill from leishmaniasis. Definitions for critical illness from leishmaniasis included the spleen reaching to the pelvic crest, haemoglobin concentration < 50 g/l, white cell count < 2 × 109/l,3 platelet count < 80 × 109 /l,3 aspartate aminotransferase concentration > 4 × upper limit of normal, serum albumin concentration < 20 g/l, and urine urea or creatinine concentration > 2 × upper limit of normal |
| Sundar-2000b | Male and female patients of any age were eligible for the study if they had typical symptoms and signs of kala-azar (e.g., fever, weight loss, hepatosplenomegaly, and pancytopenia), if they had characteristic amastigotes observed microscopically on splenic or bone marrow aspirate smears, and if they had not received prior antileishmanial treatment.  Patients were excluded from this study if they had a serious underlying illness (e.g., congestive heart failure, renal dysfunction, or diabetes) or an associated infection (e.g., tuberculosis or malaria), if they were receiving immunosuppressive therapy, or if one of the following laboratory abnormalities was present: a serum glutamic oxaloacetic transaminase value >5 times the normal value, a serum creatinine level >2.0 mg/dL, or a prothrombin time >5 seconds above the control value. |
| Sundar-2000a | Patients were eligible for the study if they (a) had symptoms or signs of visceral leishmaniasis (fever, weight loss, splenomegaly), (b) had amastigotes demonstrated on splenic aspirate smear, and (c) had not received antileishmanial therapy in the previous 3 months. Patients were excluded if they were aged <5 or >65 years, had a history of cardiac disease, were pregnant, had a serious concurrent infection such as tuberculosis or a serious bacterial pneumonia, or if 1 of the following laboratory abnormalities was present: granulocyte count: granulocyte count< 1000/mm3, haemoglobin <6 g/dL, platelet <50 000/mm3, prothrombin time >4 s above control values, bilirubin >2.0 mg/dL, serum glutamic oxaloacetic transaminase (SGOT) >200 IU, or creatinine >2.0 mg/dL. Together, these criteria would probably acetic transaminase (SGOT) >200 IU, or creatinine >2.0 mg/dL.  None of the 70 enrolled subjects was HIV seropositive. |
| Thakur-2000b | Patients aged 6-50 years with symptoms and signs suggestive of visceral leishmaniasis and aspirates of spleen or bone marrow positive for leishmania amastigotes were eligible for inclusion in the study if they gave signed informed consent. Exclusion criteria were a known allergy to aminoglycosides, treatment in the previous 12 months with a drug with recognised or presumed anti-leishmanial action, serious concomitant diseases, pregnancy or lactation (women underwent a pregnancy test at initial assessment), failure to agree to return for all follow up evaluations, and being critically ill from leishmaniasis. Definitions for critical illness from leishmaniasis included the spleen reaching to the pelvic crest, haemoglobin concentration < 50 g/l, white cell count < 2 × 109 /l3, platelet count < 80 × 109 /l3, aspartate aminotransferase concentration > 4 × upper limit of normal, serum albumin concentration < 20 g/l, and urine urea or creatinine concentration > 2 × upper limit of normal. |
| Veeken-2000 | The following clinical case definition was used: patients with fever for more than two weeks (with exclusion of malaria), in combination with either splenomegaly or wasting. In cases meeting the case definition, kala-azar was confirmed by a ⱖ 1:6400 titre of antibodies against Leishmania by the Direct Agglutination Test (and a subsequent response to antimonial treatment); or by demonstration of Leishmania on aspirates of spleen or lymph node. Patients who had received any treatment for kala-azar in the past were excluded. Nomadic patients, in whom follow up would be impossible, were excluded. Informed written consent was given by the patient or their guardian/parent. Participation in the study was voluntary, and those who did not participate were treated with Pentostam. Patients were not excluded from treatment on the basis of severe disease. Malnourished patients received supplemental or therapeutic feeding according to severity. |
| Villanueva-2000 | Concomitant infections were present in 11 patients: tuberculosis (4); Pneumocystis carinii pneumonia (PCP) (2); PCP plus esophageal candidiasis (1); PCP plus tuberculosis and Kaposi sarcoma (1); pneumococcal pneumonia (1); Escherichia coli bacteremia (1); and sinusitis (1). Inclusion: -Patients diagnosed with VL and HIV-1 coinfection between November 1996 and February 1999. -Patients were considered positive for HIV infection if they had both a positive enzyme immunoassay and Western blot assay for HIV-1. -Visceral leishmaniasis was diagnosed when amastigotes were identified in bone marrow aspirate smears on Giemsa staining or when growth of promastigotes occurred in bone marrow samples cultured in Novy-McNeal-Nicolle (NNN) medium. |
| Thakur-2001a | HIV positive cases, cases complicated with tuberculosis and renal, liver and heart diseases, patients unable to follow the protocol in all phases of study, pregnant and lactating women, refusal to give informed consent and cases previously treated with Ambisome were excluded. Certain precautions were taken in both the groups before starting treatment. The treatment was started only when the haemoglobin was above 5 dg:dl, serum electrolytes deficiency, if any, was corrected [22], and myocardial damage assessed by ECG changes caused by prior treatment with sodium antimony gluconate stabilised [23]. It was decided to withdraw patients from the trial if they developed serious side effects. |
| Moore-2001 | It may be noted that patients were included even if they had severe kala-azar, or comorbidity such as anaemia or cachexia. Pregnancy, breastfeeding, advanced age and infancy were not reasons for exclusion. Patients with a past history of kala-azar or who had received any antimonials in the past were excluded. Patients who were clinically suffering from kala-azar but had both a negative DAT test and a negative splenic aspirate were also excluded. |
| Ritmeijer-2001 | Patients previously treated for VL were excluded. Participation in the study, including HIV testing, was voluntary and patients would receive treatment with Pentostam if they were to decline.HIV status was tested with 2 rapid tests. HIV testing was not possible for all patients: some patients died before blood was drawn, and during one period of the study no test kits were available. Patients were identified using a **modified WHO clinical case definition (WHO, 1996; MSF, 1999)** of fever for >2 weeks, with exclusion of malaria, in combination with either splenomegaly or wasting. In cases meeting the case definition, VL was confirmed by a high titre Leishmaniu Direct Agglutina- tion Test (DAT; titre 2 1 : 6400). In cases with a borderline (1 : 800- 1 : 6400) or negative DAT (~1 : 800), splenic aspiration was performed. Slides of splenic aspirates were checked by an independent microscopist at a later date. Given the harsh field conditions with failures in the cold chain and DAT antigen supply, the clinicians sometimes treated patients on clinical grounds, without parasitological or DAT confirmation. |
| Thakur-2001b | Patients with renal and cardiac complication were excluded. |
| Dietze-2001 | The exclusion criteria included clinical contraindication to splenic aspiration, any history of anti-Leishmania therapy, evidence of serious underlying disease (cardiac, renal, hepatic, or pulmonary), including serious infection other than VL, acquired immunodeficiency syndrome or antibody to human immunodeficiency virus (HIV), severe protein and/ or caloric malnutrition (kwashiorkor or marasmus), glucose- 6 phosphate dehydrogenase deficiency, pregnancy, haemoglobin concentration < 5 g/100 mL, white blood cell count <1,000, platelet count < 30,000/mm3 , and a significant (> 3 times upper limit of normal) deviation in serum chemistries such as blood urea nitrogen, creatinine, alanine aminotransferase, and aspartate aminotransferase. |
| Sundar-2001 | Patients of any age with visceral leishmaniasis caused by L donovani were eligible if they had symptoms or signs of the disease (fever, weight loss, splenomegaly) and parasites were found by micros- copy in a splenic aspirate smear.Patients were excluded if they were pregnant or breast feeding, HIV positive, had a serious concurrent infection such as tuberculosis or bacterial pneumonia, or if they had a granulocyte count < 1×109/l, haemoglobin concentration < 40 g/l, or platelet count < 40×109/l. Eight patients were excluded by these criteria. |
| Das-2001 | Inclusion: VL patients unresponsive to antimony. Those patients who were **critically ill,** aged below five years and above 60 years, hemoglobin concentration below 40 g/1, having concurrent TB, renal impairment, cardiac disease or jaundice were excluded from the trial. |
| Sundar-2002 | Potentially eligible patients were 12 years of age or older with visceral leishmaniasis suspected on the basis of clinical presentation (fever, splenomegaly, and cytopenia) and diagnosed by the presence of leishmania in splenic aspirates). Criteria for exclusion were a platelet count below 50,000 per cubic millimeter, a white-cell count below 1000 per cubic millimeter, a hemoglobin concentration of less than 6 g per deciliter, results on liver-function tests (serum aspartate aminotransferase and alanine aminotransferase concentrations) more than three times the upper limit of normal, a bilirubin concentration more than twice the upper limit of normal, serum creatinine or blood urea nitrogen values more than 1.5 times the upper limit of normal, other major medical illness including human immunodeficiency virus infection or severe malaria, pregnancy or lactation, inability to maintain use of contraception for the period of treatment plus two months, and previous therapy with amphotericin B. |
| Sundar-2002a | Patients of any age or sex were eligible for enrolment into the trial if they had signs and symptoms of VL confirmed by the presence of parasites in splenic or marrow smears if they had failed to respond or VL relapsed after a full course of Sbv treatment. Pregnant or lactating women, HIV-positive patients, and intravenous drug abusers were excluded from the trial. |
| Laguna-2003 | Exclusion criteria included patients with pancreatitis, prothrombin activity <40%, aminotransferase levels 10× the upper normal limit, myocardiopathy, heart failure, a Qt corrected interval >500 ms, creatinine levels >twice the upper normal limit, allergy to either ABLC or meglumine antimoniate, concomitant treatment with dideoxy- cytidine or dideoxyinosine and a life expectancy of <6 months. Women of childbearing potential were excluded if they were pregnant, might become pregnant, or were lactating. Active opportunistic infections were not an exclusion criterion. |
| Sundar-2003b | A total of 203 patients participated. Patients of all ages and both sexes were potentially eligible if they had symptoms and signs suggestive of VL (i.e., fever with chills, rigor, and splenomegaly) with demonstrable Leishmania parasites in splenic or bone marrow aspirate smears. Pregnant and lactating women, HIV-positive individuals, and individuals receiving concomitant antileishmanial drugs were excluded from the study. Patients were considered to have prior treatment failure if they had received adequate antileishmanial treatment, consisting of either sodium antimony gluconate (SAG; also known as sodium stibogluconate) at a dosage of 20 mg/kg for 30 days or 15 infusions of AmB administered at a dosage of >= 0.75 mg/kg. |
| Figueras Nadal-2003 | Children between 0 and 14 years old diagnosed with visceral leishmaniosis through direct visualization of Leishmania amastigotes by Giemsa staining OR presence of Leishmania promastigotes in bone marrow aspirate culture OR detection of antibodies through indirect immunofluorescence (>1/80) OR ELISA analysis (>0,80). Exclusion: a) Previous or concomitant treatment with amphotericin B, antimonials derivatives or other drug used for the treatment of visceral leishmaniasis in the year prior to the enrolment in the study; b) Immunosuppression. |
| Sundar-2003 | Children between the ages of 2 to 11 years with signs and symptoms suggestive of VL and demonstrable parasites in splenic aspirates were screened for eligibility in the study. Those with hemoglobin <6 g/dl, leukocyte count <2000/nl, platelet count <50 000/ nl, blood urea nitrogen (BUN) or serum creatinine >1.5 times of upper normal limit and alanine aminotransferase (ALT) >3 times upper limit were excluded from the study. Patients with HIV positivity or any concomitant non-compensated renal, hepatic, malignant or infectious diseases were excluded. Because ophthalmologic evaluation was to be done, children with retinal diseases were also excluded. |
| Syriopoulou-2003 | Eligible participants were children up to 14 years of age who had clinical features that were consistent with active VL (e.g., fever, splenomegaly, anaemia, leukopenia, or thrombocytopenia) and who had visible parasites or amplified Leishmania DNA detected on bone marrow aspirates. Children with concomitant disease, known allergy to study compounds, known HIV seropositivity, or other immunosuppression, and children who had previously received treatment for leishmaniasis were excluded from the study. |
| Rijal-2003 | Parasitologically proven VL cases with no history of previous treatment with SSG were included after obtaining informed consent from the patient or his/her guardian. Only patients from the 3 neighbouring districts of Sunsari, Morang, and Saptari were included as follow-up would not be practically possible for patients coming from more remote districts. There was no other exclusion criteria. All the patients were negative to HIV testing |
| Thakur-2004a | Only patients who provided informed consent (or that of their guardians, if they were children) were enrolled. Patients were excluded if they had <6.0 g haemoglobin/dl blood, complications such as pneumonia, jaundice, tuberculosis, renal or cardiac disease, diabetes or HIV infection, serum concentrations of aspartate and/or alanine aminotransferase that were at least three times the upper limit of normal, a serum concentration of creatinine that was more than 1.5 times the upper limit of normal or had already received treatment with antimony, pentamidine, AMB or any other antileishmanial drug. Patients found to be infected with hookworm, Ascaris and/or Entamoeba histolytica/E. dispar were not excluded, as these parasites are very common in the study area but were treated appropriately (concurrently with antileishmanial treatment). |
| Sundar-2004 | Patients were eligible if they had symptoms and signs of VL (fever, weight loss, and splenomegaly) and if microscopic analysis of a splenic aspirate smear revealed parasites  Patients were excluded if they were pregnant or breast-feeding, were HIV seropositive, or had a serious concurrent infection, such as tuberculosis or bacterial pneumonia. Exclusion criteria also included a granulocyte count of <1000 granulocytes/mm3, a hemoglobin level of <3.5g/dL, or a platelet count <40,000 platelets/mm3; hepatic transaminase or total bilirubin levels that were >3 times the upper limit of normal; a serum creatinine level of 12.0 mg/dL; or a prothrombin time of >5 s above the control time |
| Thakur-2004b | Patients positive for HIV and other co-infections (n=28) were excluded. |
| Bhattacharya-2004 | Potentially eligible patients were 2–11 years of age, were of either sex, and had VL suspected on the basis of clinical presentation (fever, splenomegaly determined by palpation, anemia, and cytopenia) and confirmed by the demonstration of amastigotes in splenic aspirate specimens. Exclusion criteria were values for the formed elements of the blood suggesting a premorbid state and severe disease (platelet count, <50,000 platelets/mL; leukocyte count, <1000 leukocytes/mL; hemoglobin concentration, <6 g/dL; serum aspartate aminotransferase (ASAT) or alanine aminotransferase levels of >3 times the upper limit of normal; bilirubin level of more than twice the upper limit of normal; and serum creatinine or blood urea nitrogen levels of >1.5 times the upper limit of normal), documentation of other serious coincidental medical illnesses, HIV seropositivity, and, for patients capable of reproduction, pregnancy |
| Wasunna-2005 | Subjects eligible for inclusion had a clinical diagnosis of VL with symptomatic disease and the diagnosis confirmed by the presence of Leishmania amastigotes in splenic aspirates. Subjects 5−65 years of age of either sex were included in the study.  Females that were pregnant or lactating were excluded. Additional exclusion criteria included known hypersensitivity to sitamaquine, use of an anti-leishmanial agent within 10 days or of an investigational compound within 30 days (or 5 half-lives) of the start of the study, subjects with marked deviations in serum chemistry or abnormal hematologic markers outside those expected to be caused by VL, patients with serious underlying disease, including human immunodeficiency virus (HIV) infection, malnutrition, or **severe kala-azar,** glucose-6-phosphate dehydrogenase deficiency, or previous inclusion in this study. |
| Jha-2005 | Subjects aged 5−65 years of either sex were included in the study. Females of child-bearing age had to have a negative pregnancy test and agree to practice effective contraception throughout the study. Exclusion criteria included known hypersensitivity to sitamaquine, receipt of an antileishmanial agent within 10 days or of an investigational compound within 30 days (or 5 half-lives) of study start, sub- jects with marked deviations in serum chemistry, patients with serious underlying disease, including HIV infection, mal- nutrition or severe kala-azar, pregnancy or lactation, G6PD deficiency, or previous inclusion in this study. |
| Das-2005 | All patients attending the outpatient department of our institution with fever, splenomegaly, Leishman-Donovan (LD) bodies in the bone marrow and/or splenic aspirate and without any history of treatment for kala-azar were included in the study. Exclusion: Not reported |
| Ritmeijer-2006 | Males aged >=15 years with parasitologically and/ or serologically confirmed VL attending Humera Hospital and Mycadra Health Center were enrolled in the study. Because of the potential teratogenicity of miltefosine, females were excluded. Previous antileishmanial treatment was recorded at hospital admission. Patients were enrolled in the study after giving informed consent. Potentially eligible patients were only excluded if they had such severe comorbidity that they were considered to be likely to die during the month’s treatment. Patients with previous antileishmanial treatment were only admitted if they had a positive aspirate result.  The World Health Organization case definition of VL was used for initial screening: a history of fever for >2 weeks (with malaria excluded) in **combination with wasting,** and either splenomegaly or lymphadenopathy; A total of 580 adult male VL patients were enrolled in the study |
| Sundar-2006 | Patients were eligible for inclusion in the study if they had signs and symptoms of active VL (i.e., fever and splenomegaly) and if their splenic aspirate smear showed characteristic Leishmania amastigotes. Pregnant or lactating female subjects and patients who were found to be HIV positive by serological testing were excluded from the study. Patients who either were critically ill or had a concurrent serious infection/ illness also were excluded. Also excluded were patients who had a granulocyte count of <1000 granulocytes/mm3, a hemoglobin level of <3.5 g/dL, or a platelet count of<40,000 platelets/mm3; hepatic transaminase or total bilirubin levels that were 3 times the upper limit of the range considered to be normal; a serum creatinine level of 12.0 mg/dL, or a prothrombin time 15s above the control time. |
| Singh-2006 | Children, who had received less than 30 days course of SAG, with bleeding diathesis, liver disorder (ALT & AST>3 times, serum bilirubin >2 times of normal), renal dysfunction (BUN and serum creatinine; 1.5 times of normal), co-existing malaria or HIV, neutrophil count less than 1000/cu mm and platelet count less than 40,000/cu mm were excluded from study. |
| Singh-2006 | Children with bleeding diathesis, liver disorder (ALT & AST>3 times, serum bilirubin >2 times of normal), renal dysfunction (BUN and serum creatinine; 1.5 times of normal), co-existing malaria or HIV, neutrophil count less than 1000/cu mm and platelet count less than 40,000/cu mm were excluded from study. Patients who had received incomplete course of sodium stibogluconate (SAG) were also excluded from study. |
| Sundar-2007b | Pregnant or breast-feeding women, individuals who were seropositive for HIV, and individuals with a serious concurrent infection (e.g., tuberculosis or bacterial pneumonia) were excluded from the study. Exclusion criteria also included the following findings: a leukocyte count of <1000 cells/mm^3^ , a hemoglobin concentration of<3.5 g/dL, or a platelet count of <40,000 platelets/mm3 ; hepatic transaminase or total bilirubin levels of >3 times the normal level; serum creatinine level, 12.0 mg/dL; and prothrombin time, >5 s greater than the control level. |
| Sundar-2007a | Inclusion criteria were parasitologically positive splenic or bone marrow smear; negative serologic testing for the human immunodeficiency virus (HIV); hemoglobin level of at least 5.0 g per deciliter; white blood count greater than or equal to 1×10exp9 per liter; platelet count greater than or equal to 50×10exp9 per liter; levels of aspartate aminotransferase, alanine aminotransferase, and alkaline phosphatase less than or equal to three times the upper limit of the normal range; prothrombin time less than or equal to 5 seconds greater than that among control subjects; and serum creatinine and potassium levels within the normal limits.  Exclusion criteria were treatment for visceral leishmaniasis during the 2 weeks before enrolment, a hearing loss of 75 dB in frequencies 1 through 8 kHz, a history of vestibular or auditory dysfunction, prior treatment with amphotericin without response, allergy or hypersensitivity to aminoglycosides, significant proteinuria (≥2+ on strip testing), significant coexisting diseases possibly affecting the response to the study treatment response, and pregnancy or lactation |
| Bhattacharya-2007 | Eligible patients were of either sex between 2 and 65 years of age with a clinical diagnosis of active VL (e.g., fever or splenomegaly) and diagnosis confirmed by splenic aspirate showing characteristic amastigotes. Exclusion criteria were ongoing pregnancy or breastfeeding, refusal to use contraceptive methods during the treatment period and 2 months thereafter (as advised by the clinical investigators), HIV-positive serology, or any condition or situation compromising with compliance to study procedures.Approximately 10% of patients did not fill eligibility criteria for inclusion in the study for various reasons (e.g., they had malaria; other causes of fever-like tuberculosis, typhoid fever, or urinary tract infection, etc.; or cirrhosis of liver or severe anemia [hemoglobin level >=4 g/dL]).  Patients with leukocyte count >1000/mm3 and hemoglobin level >=4 g/dL were included |
| Mueller-2007 | Inclusion: Unresponsiveness to AmBisome in some Sudanese patients with kala-azar. Exclusion: Not clear |
| Sundar-2008b | Patients >=12 years of age were eligible for the study if they had symptoms and signs of kala-azar (e.g., fever, weight loss, and splenomegaly) and parasites demonstrated by microscopic examination of splenic aspirate smear Pregnant or breast-feeding women and individuals who were seropositive for HIV or who had a serious concurrent infection, such as tuberculosis or bacterial pneumonia, were excluded. Exclusion criteria also included granulocyte count !1000 cells/mL, hemoglobin level >3.5 g/dL or platelet count >40,000 platelets/mL, hepatic transaminase levels or total bilirubin >3 times the upper limit of normal, serum creatinine level >2.0 mg/dL, and prothrombin time 15 sec above control. If they were randomized to receive miltefosine, women of child-bearing age with negative pregnancy test results were counselled about the potential teratogenic effects of miltefosine and were offered contraception in the form of a depot preparation of progesterone. All such women enrolled in this study gave consent and received the injection |
| Sundar-2008a | Patients of either sex aged 12–65 years (both inclusive) with signs and symptoms of VL (fever, weight loss and splenomegaly), and demonstrable parasite in splenic smears were included in this study. Patients who had failed to respond to sodium stibogluconate treatment previously were also included after a washout period of 10 days  Patients were excluded if they were breast feeding or pregnant, if they were HIV positive or if they had previously failed treatment with amphotericin B for VL. Additional exclusion criteria were a granulocyte count of <1000 mm)3; a haemoglobin level <6 gm ⁄dl or a platelet count <50 000 mm)3; hepatic transaminases >3 times the normal upper limit and ⁄or total bilirubin >2 times of normal upper limit; serum creatinine ‡ 1.5 times of normal upper limit; or a prothrombin time >5 s above the control range. Patients with concomitant life-threatening disease or serious concurrent infection such as tuberculosis or bacterial pneumonia and those allergic to amphotericin B or any other ingredients of the emulsion formulation were also excluded from the study |
| Thakur-2008 | Patients were excluded if they had a haemoglobin concentration below 50g/dl, had complications such as pneumonia, jaundice, tuberculosis, renal and cardiac diseases, or had received treatment with antimony or amphotericin B (AMB) or any treatment for kala-azar or refused to be included in the study. Patients with parasitological infections with hookworm, roundworm and Entamoeba histolytica were not excluded and these infections were treated concurrently |
| Mueller-2008 | Twenty-five (11.9%) patients with malaria, 22 (10.5%) with respiratory tract infections, and 21 (10.0%) with other bacterial infections (such as typhoid fever, otitis media, pharyngitis, brucellosis or dysentery) were given specific treatments for their accompanying infections |
| Sundar-2009a | Patients of either sex and 18–65 years of age (both inclusive) with signs and symptoms of VL (fever, weight loss, and splenomegaly) who had demonstrable parasites in splenic smears were included in this study. Patients who had failed treatment with antileishmanial drugs other than amphotericin B were also included after completion of a washout period of 10 days.  Patients were excluded if they were breast feeding or pregnant, and had positive serologic results for infection with human immunodeficiency virus (HIV). Exclusion criterion also included a granulocyte count < 1,000 cells/mm 3; a hemoglobin level < 6 g/dL, a platelet count < 50,000 cells/mm 3; levels of hepatic aminotransferases> 2.5 times the upper limits of normal levels, total bilirubin levels > 1.5 times the upper limit of normal ranges; pro- thrombin time > 5 seconds than that of controls, and serum creatinine levels ≥ 1.5 times the upper limit of normal ranges. Patients with concomitant life-threatening disease or serious concurrent infection such as malaria, tuberculosis, or bacterial pneumonia, and those allergic to amphotericin B or any other ingredients in the emulsion formulation were also excluded from the study |
| Das-2009 | Patients of either sex aged between six to 60 years unresponsive to SSG after a full course of treatment and having com- plaints of continued fever with rigor, hepato-splenomegaly, anorexia, anemia, and loss of weight, sputum for acid-fast bacilli (AFB) and chest x-ray (PA-view) negative for tuberculosis or pneumonia, and demonstration of parasite in bone marrow (BM)/splenic(SPL) aspiration were the inclusion criteria.  The confirmed VL cases with partial dosages of SSG and with associated diseases like tuberculosis (TB), HIV/AIDS, and malaria, pregnant women, lactating mother, and diabetics were not included in the study. The individuals who had past history of Kala-azar in last five years and having Hb< 6 g/dl, platelet count <50,000/mm3 with significant abnormalities in liver and renal function tests were the exclusion criteria. |
| Sundar-2009b | Patients of either sex who were 5–55 years of age with signs and symptoms of VL (ie, fever, weight loss, and splenomegaly) who had demonstrable parasites in splenic smears were included in this study. Patients with relapsed VL or who experienced treatment failure while receiving a regimen that did not contain paromomycin were also included. Women with child-bearing potential were excluded from the study if they were pregnant, or were not using any contraception. Exclusion criterion included a granulocyte count of less than 1000 cells/mm3, a hemoglobin level of less than 4 g/dL, a platelet count less than 40,000 platelets/ mm3, liver function test results greater than 3 times of upper limit of normal, normal, prothrombin time 5 sec more than that for control subjects, serum creatinine level above the upper limit of normal, abnormal audiometric, and/or vestibular function, serological test results positive for HIV, and concomitant severe medical conditions. Patients who were allergic to aminoglycosides, those treated with a parenteral aminoglycoside within 28 days prior to randomization or with paromomycin at any time and those who had taken VL treatment within the past 14 days were excluded. |
| Adam-2009 | Pregnant women with symptoms and signs suggestive of VL, such as fever, weight loss, anemia, and hepatosplenomegaly, were approached to participate in the study. Besides being pregnant, the inclusion criteria were confirmation of Leishmania infection by the presence of amastigotes in Giemsa-stained smears of bone marrow, and signing an informed written consent form. None of the patients had malaria or HIV |
| Shahian-2009 | No patient had immunosuppression caused by HIV infection or renal transplantation and none of them had received medications inducing pancreatitis or prior antileishmanial therapy |
| Hailu-2010 | Patients were excluded if they: (1) had taken any antileishmanial drug in the preceding 6 months; (2) showed severe protein or caloric malnutrition (Kwashiokor or marasmus); (3) had previous hypersensitivity reaction to SSG or aminoglycosides; (4) had a concomitant severe infection (except HIV) or any other serious underlying (cardiac, renal, hepatic) disease; (5) had conditions associated with splenomegaly, such as schistosomiasis; (6) had a history of cardiac arrhythmia or an abnormal electrocardiogram (ECG); (7) were pregnant or lactating; (8) had any relevant outliers of safety laboratory parameters (hemoglobin ,5 g/dL, white blood cells ,16103/mm3, platelets ,40,000/mm3, liver function test values .3 times higher than upper limit of normal, serum creatinine values above upper limit of normal); or (9) had clinical hearing loss. HIV infection was not an exclusion criterion Eligible patients were 4-60 years old and had to have clinical symptoms (fever and splenomegaly) and a diagnosis of VL, confirmed by microscopic verification of parasites in spleen, lymph node or bone marrow tissue aspirates. |
| Thakur-2010 | Exclusion criteria included patients who had tuberculosis, infection with human immunodeficiency virus, acquired immunodeficiency syndrome, kidney and heart disease, and leukocyte counts < 1,000 cells/μL, a hemoglobin concentration < 5 g/dL, serum aspartate aminotransferase and alanine aminotransferase levels > 3 times the upper limit of the reference range, thrombocyte counts < 60,000 cells/μL, or serum creatinine levels > 1.5 times the upper limit of the reference range. If patients had hemoglobin levels < 5 g/dL or a thrombocyte count < 65,000 cells/μL, whole blood transfusions or platelet transfusions were given, respectively. If these two parameters reached acceptable levels, only then were the patients included in the trial. |
| Musa-2010 | Patients were excluded from the study if they: had negative bone-marrow smears; were clinically contraindicated to having a bone-marrow aspirate; received any anti-leishmania drug in the past 6 months; had severe protein or caloric malnutrition (Kwashiorkor or marasmus); had previous hypersensitivity reaction to aminoglycosides; suffered from a concomitant severe infection, ie tuberculosis, HIV, or any other serious underlying disease (cardiac, renal, hepatic); suffered from other conditions associated with splenomegaly such as schistosomiasis; had previous history of cardiac arrhythmia or an abnormal electrocardiogram (ECG); were pregnant or lactating; or had pre-existing clinical hearing loss. If tuberculosis or schistosomiasis were suspected, these were screened through laboratory testing. Additionally, patients with the following laboratory values were excluded: hemoglobin less than 5 g/dL; white blood cell less than 103/mm3; platelets less than 40,000/mm3; liver function test values more than three times the normal range; and serum creatinine values outside the normal range for age and gender. |
| Sundar-2010 | Patients between the ages of 2 and 65 years were eligible if they had symptoms and signs of leishmaniasis (e.g., fever, weight loss, and splenomegaly) and if parasites were shown on microscopy of a splenic aspirate smear.  Patients who were seropositive for the human immunodeficiency virus (HIV) or who had a serious concurrent infection, such as tuberculosis or bacterial pneumonia, were excluded. Exclusion criteria also included a white-cell count of less than 750 percubic millimeter, a hemoglobin level of less than 3.5 g per deciliter, and a platelet count under 40,000 per cubic millimeter;levels of hepatic aspartate aminotransferase or total bilirubin of more than five times the normal range; a serum creatinine level of more than 1.5 times the upper limit of the normal range; and a prothrombin time of more than 4 seconds above the control level. |
| Mondal-2010 | Based on the inclusion exclusion criteria (Protocol S1): patients of all ages and both sexes were potentially eligible if they presented the clinical symptoms of prolonged fever, hepatosplenomegaly, and were confirmed to be VL by K39 strip test and detection of Leishmania parasites in the splenic or bone marrow aspirate.  - Patients of all ages and both sexes -Consistent signs and symptoms of active VL. -Confirmed diagnosis of VL with positive identification of parasite from bone marrow or splenic aspirate or K39 strip test. -Confirmed VL diagnosis patients fully informed consent (self or parent or authorized relative) of their willingness to undergo treatment with the new drug (Fungisome) at the specific dose. -Patients willing to participate in all treatment and follow-up visits regularly at monthly intervals for six months at the out-patient department at the school of Tropical Medicine    Excluded: -HIV-positive individuals with VL -VL Patients already receiving antileishmanial drugs will be excluded from the study. -Patients who have hypersensitivity to the drug or its constituents. -Patients who have associated disease altering liver function tests. -Pregnant women. |
| Rijal-2010 | Parasitologically proven kala-azar cases who were treatment-naïve to Sbv therapy were enrolled in the study after obtaining informed consent.  Exclusion: Not presented |
| Singh-2010 | Children with bleeding diathesis, liver disorder [ala- nine aminotransferase (ALT) and aspartate amino- transferase (AST) >3 times, serum bilirubin >2 times of normal], renal dysfunction [Blood urea nitrogen (BUN) and serum creatinine, 1.5 times of normal], co-existing malaria or HIV, neutrophil count <1000mm-3 and platelet count less than 40 000mm-3 were excluded from study. Patients who had received incomplete course of SAG were also excluded from study |
| Sinha-2010 | Inclusion: (1) clinical symptoms consistent with kala-azar, (2) ≥ 2 years of age, (3) residence within Vaishali District, (4) provision of written informed consent, and (5) a positive rK39 rapid diagnostic test (DiaMed-IT-Leish; in northeast India, rk39 has shown 99% sensitivity and 100% specificity with whole-blood samples) or a parasitological diagnostic test result.  Exclusion: Patients excluded from the study were those with (1) post-kala-azar dermal leishmaniasis, (2) prior treatment of current kala-azar infection with ≥ 20 mg/kg of liposomal amphotericin B, (3) known allergic reaction to amphotericin B, or (4) human immunodeficiency virus, tuberculosis, or malaria co-infections.  Pregnancy was noted for 3 of 46 (6.5%) women of childbearing age. |
| Sundar-2011b | Reasons for exclusion were haemoglobin concentrations less than 50 g/L, total leucocyte count less than 1×10⁹/L, serum creatinine concentration outside the normal range of 55–140 μmol/L, platelet count less than 40×10⁹/L, serum aminotransferase concentration higher than three times the upper limit of the normal ranges, bilirubin concentration more than 34·2 μmol/L, prothrombin time more than 5 s longer than control, positive serology for HIV or hepatitis B or C viruses, severe concurrent illness, and receipt of any antileishmanial or antifungal drug in the previous 45 days. Pregnant and breast-feeding women, patients with known hypersensitivity to the study drugs, and those with diabetes, hypertension, or tuberculosis were also excluded. |
| Sundar-2011c | Eligible patients were men or women 16−50 years of age with VL symptoms or signs and confirmed VL  Exclusion criteria were renal, hepatic or biliary disease; renal or hepatic impairment; cardiac disease, arrhythmia, or conduction abnormalities; clinically relevant electrocardiogram (ECG) results or laboratory values; serious underlying disease or infection; glucose-6-phosphate dehydrogenase deficiency (based on phenotype testing); positive results for antibodies against human immunodeficiency virus, hepatitis B surface antigen, or antibodies to hepatitis C virus; contraindication to splenic or bone marrow aspiration; hypersensitivity to study treatments; treatment with an established anti-leishmania drug within 30 days or 5 half-lives (whichever was longer) of the start of the study; or treatment with prohibited medication. Pregnant or nursing women were excluded; a negative urine pregnancy test result was required from female patients at screening and before dosing, plus an agreement to use contraception for two weeks |
| Sundar-2011a | Patients of age 2–65 yrs with mild-moderate kala-azar; lack of concomitant diseases such as malaria, HIV, and tuberculosis; lack of pregnancy or lactation; no previous treatment with antimony or paromomycin unless the treatment terminated two months prior to the study and the patient was worsening |
| Rahman-2011 | Patients were 2–65 years of age. Both male and female patients participated, and patents were enrolled during October 2006–September 2007. Inclusion criteria were signs and symptoms compatible with VL (fever for at least two weeks, palpable splenomegaly, weight loss by history), positive serologic test result for leishmaniasis (rK39), hemoglobin level ≥ 6 g/dL, no infection with HIV, not pregnant or lactating, not being currently treated with an antileishmanial compound, and no significant concomitant medical condition. Malaria and enteric fever were ruled out by blood smears and the Widal test, respectively. If present, malaria and enteric fever were treated. These patients could then be admitted into the study. If tuberculosis was suspected on clinical grounds, the patient was not eligible for this study. |
| Sinha-2011 | Exclusion criteria included concurrent illness such as HIV, malaria, and tuberculosis; a history of hearing loss that could confound clinical detection of potential ototoxicity; recent or current exposure to medications that could result in com- pounded toxicities. Before study enrolment, informed consent was obtained from every patient or a legally authorized representative |
| Sudarshan-2011 | Forty-six patients with parasitologically confirmed VL (6–55 years, males and females, and HIV negative) were enrolled in the study. Confirmation of VL was done by demonstration of amastigotes in Giemsa-stained smears of splenic aspirate. |
| Sundar-2012 | Criteria for exclusion were current pregnancy; current breast-feeding; seropositivity for human immunodeficiency virus; presence of a serious illness or concurrent infectious disease, such as tuberculosis or bacterial pneumonia; granulocyte count <1000 granulo- cytes/mm3; hemoglobin level <5.0 g/dL; platelet count <40 000 platelets/mm3; hepatic transaminase levels >5 the normal level; total bilirubin level >2.0 mg/dL; serum creatinine level above the upper normal limit; prothrombin time >5 seconds above control; and/or inability of the subject or guardian to provide written informed consent.  Because of the long half-life of miltefosine and its known teratogenic potential, women of childbearing age were asked to practice contracep- tion during treatment and for an additional 3 months after completion of treatment. Treatment was directly observed by a clinic nurse, who observed the swallowing of the pills |
| Musa-2012 | Briefly, patients aged 4–60 years with parasitologically confirmed VL were included, but patients with **very severe VL or those with contraindications were excluded**  Being HIV-positive was not an exclusion criteria but the original protocol stated that there was to be a sufficient number of patients for a subgroup analysis excluding HIV patients (if deemed necessary).  Patients were excluded if they: (1) had taken any antileishmanial drug in the preceding 6 months; (2) showed severe protein or caloric malnutrition (Kwashiokor or marasmus); (3) had previous hypersensitivity reaction to SSG or aminoglycosides; (4) had a concomitant severe infection (except HIV) or any other serious underlying (cardiac, renal, hepatic) disease; (5) had conditions associated with splenomegaly, such as schistosomiasis; (6) had a history of cardiac arrhythmia or an abnormal electrocardiogram (ECG); (7) were pregnant or lactating; (8) had any relevant outliers of safety laboratory parameters (hemoglobin <5 g/dL, white blood cells <1103/mm3, platelets <40,000/mm3, liver function test values >3 times higher than upper limit of normal, serum creatinine values above upper limit of normal); or (9) had clinical hearing loss. |
| Patra-2012 | Patients with following were excluded from the study: platelet count < 50,000 /mm 3 ; total leukocyte count < 1000 /mm 3 ; hemoglobin concentration < 6 g/dL; bilirubin more than twice the upper limit of normal and serum creatinine or blood urea nitrogen levels of 1.5 times the upper limit of normal. No patients had been treated previously for VL. Cases with other serious concurrent medical illness, HIV seropositivity, and pregnancy were also excluded |
| Rijal-2013 | Exclusion criteria included pregnancy or breast feeding, as well as women of child-bearing age who refuse or are unable to maintain contraception for a period of two months after completion of treatment. Also SGPT/SGOT > 3 times, serum bilirubin > 2 times and serum creatinine> 1 5 times the upper limit of normal values, and cases with severe anemia or known kidney or liver disease were excluded. The patients that did not match the inclusion criteria were treated with amphotericin B (1 mg/kg/day for 14 days). MIL relapse cases were also not included. |
| Khalil-2014 | Patients with age of at least 4 years, confirmed HIV-negative, parasitologically-confirmed non-severe VL, were enrolled in three centres   Exclusion criteria were signs/symptoms of severe VL (patients who were very weak, unable to walk, bleeding, jaundiced, suffering from sepsis and other concomitant infections/illnesses); anti-leishmanial or unlicensed investigational treatments within six months; underlying chronic disease such as severe cardiac, pulmonary, renal, or hepatic impairment; serum creatinine outside the normal range; liver function tests more than 3 times the normal range; platelet count less than 40,000/mm3; known alcohol abuse; pregnancy or lactation; concomitant acute drug usage for malaria and bacterial infection; pneumonia within last 7 days; known hypersensitivity to AmBisome or amphotericin B; any other condition which may invalidate the trial |
| Ostyn-2014 | No exclusion criteria reported. In our study area HIV co-infection is rare, HIV is tested for and when positive, patients are treated with amphotericin B, so our results on miltefosine-treatment concern an HIV-negative population. |
| Cota-2014 | Not clear |
| Sundar-2014 | Other inclusion criteria were hemoglobin (Hb) >=5 g/dL, white blood cells count >=1000/cmm, platelet count >=50000/cmm, prothrombin time <=4 seconds above the control, and alkaline transaminase, aspartate transaminase, and alkaline phosphatase <=2.5 times the upper limit of normal. Patients with past history of treatment with AmB or any other drug for VL within 30 days prior to screening, major surgery within 2 weeks prior to screening, concurrent malaria, alcoholism or illicit drug use/abuse or any condition associated with poor compliance, hypersensitivity to AmB, inactive ingredients of ABLE and LAmB formulations were excluded from the study. Patients who received any of the prohibited medications (any other investigational drugs, antileishmanial drugs other than study drug, corticosteroids, skeletal muscle relaxants, cyclosporine, digoxin, vancomycin, aminoglycosides, antifungal, immunosuppressive agents, and all potentially nephrotoxic drugs), who were positive for human immunodeficiency virus, hepatitis C virus and hepatitis B surface antigen infections and immune-compromised, were also excluded from the study. The following is from study protocol (supplemental file): Inclusion: Non-pregnant, non-lactating females of age ≥5 years, and woman of childbearing potential (any woman who has reached menarche) who are willing to use acceptable methods of contraception like long term acting injections ex. Depo-Provera. |
| Mondal-2014 | We excluded participants with a history of intercurrent or presence of clinical signs or symptoms of uncontrolled concurrent diseases or conditions before start of study treatment, any condition that might prevent the patient from completing the study therapy and subsequent follow-up (investigator assessed), a history of allergy or hypersensitivity to amphotericin B, previous treatment for visceral leishmaniasis within 2 months of enrolment, previous treatment failure with amphotericin B, post-kala-azar dermal leishmaniasis, and pregnant women. We excluded pregnant women from this study because present evidence about safety of liposomal amphotercin B during pregnancy is limited. |
| Jamil-2015 | Exclusion criteria included pregnancy or lactation; active tuberculosis or taking antituberculosis medications; previous treatment with PMIM; clinically significant anemia; current or history of clinically significant renal or hepatic dysfunction; serum creatinine above the upper limit of normal range; proteinuria; history of hepatitis B or C or HIV positive; history of hearing loss; significant coexisting disease; any history of VL or treatment for VL; history of hypersensitivity to aminoglycosides or sulfite; and concomitant use of other aminoglycosides, nephrotoxic and ototoxic drugs, or immunosuppressive drugs. |
| Sundar-2015 | Patients were excluded in the study if they had hemoglobin of < 5 g/dL, serum creatinine or blood urea nitrogen (BUN) > 1.5 times the upper limit of normal, a platelet count < 40,000/m3, serum bilirubin > 2 mg/dL, prothrombin time of more than 5 seconds above the control levels, history of VL treatment in the last 45 days, hepatitis B or C, human immunodeficiency virus (HIV), active tuberculosis or another serious illness or associated disease known to alter liver/kidney functions, or known hypersensitivity/allergy to the study drug or their constituents or were women who were pregnant or lactating |
| Goswami-2016 | 120 treatment naïve non HIV; The inclusion criteria were as follows: all patients with symptoms and signs suggestive of VL (fever for more than 2 weeks and splenomegaly) with rK39 immunochromatographic strip test (Kalazar Detect™; InBios International Inc., Seattle, WA) positivity and demonstrable amastigotes of Leishmania donovani parasites in splenic aspirates between 2008 and 2011.  The following patients were excluded from the study: pregnant and lactating women, human immunodeficiency virus-positive individuals, those previously treated with or receiving concomitant antileishmanial drugs, patients with platelet count below 50,000/?L, white blood cell count below 1,000/?L, serum aspartate aminotransferase and alanine aminotransferase levels more than three times the upper limit of normal, serum creatinine or urea values more than 1.5 times the upper limit of normal, and patients with other major medical illness such as ischemic heart disease, congestive cardiac failure, and stroke. |
| Wasunna-2016 | Eligible patients were HIV negative, and aged between 7 and 60 years with parasitologically confirmed VL who signed an informed consent (if aged 18y and over) or whom the parent or legal guardian consented to participate in the study (if under 18y). The target population was primary cases, so known relapse cases, or receipt of any anti-leishmanial drugs in the previous 6 months, was an exclusion criterion. Other exclusion criteria were: severe protein and/or caloric malnutrition defined as kwashiorkor or marasmus in children and BMI <15 in adults; previous history of hypersensitivity reaction to SSG or amphotericin B; concomitant severe infection such as TB or other serious underlying disease which would preclude evaluation of patients response to the study medication; other conditions associated with splenomegaly such as schistosomiasis; previous history of cardiac arrhythmia or an abnormal ECG; Hb<5 g/dL; WBC <103/mm3; platelets <40,000/mm3, abnormal liver function tests (ALT and AST) of more than three times the upper limit of the normal range, serum creatinine outside the normal range for age and gender, and major surgical intervention within two weeks prior to enrolment. Due to the potential teratogenicity of miltefosine, females of child bearing age were also excluded. |
| Pandey-2016 | Females and males between 6-70 years of age having signs and symptoms sug- gestive of VL (fever of 2 weeks with splenomegaly, not responding to antimalarials and antibiotics) and diagnosed for VL infection as per the prevailing guidelines of VL diagnosis in national kala-azar elimination program were included in this study.  Criteria for exclusion were pregnant/breast-feeding women, women of child-bearing age not consenting to avoid pregnancy during and after 6 months of treatment, seropositive for HIV, presence of a serious illness, concurrent diseases, such as tuberculosis or bacterial pneumonia, hemoglobin level < 5.0 g/dL, granulocyte count < 1,000 granulocytes/mm3, platelet count < 40,000 platelets/mm3, hepatic transaminase levels > 5 times of the normal limit, total bilirubin level > 2.0 mg/dL, serum creatinine level above the upper normal limit (> 1.5 mg/dL), prothrombin time > 5 seconds above control, and/or inability of the subject or guardian to provide written informed consent. |
| Borges-2017 | Eligible patients were children aged 6 months-12 years who met the inclusion criteria for VL diagnosis (presence of fever for at least two weeks associated with splenomegaly). Participation in this study was voluntary upon the signature of the informed consent form by the patients' parents or legal guardians. Treatment naïve children and adolescents with VL without signs of severe illness were treated.  Exclusion criteria were the following: patients who underwent previous treatment with leishmanicidal drugs, clinically evident jaundice (total bilirubin > 2.5mg/dL), hemorrhages with coagulation disorders, generalized edema,signs of toxemia, severe malnutrition according to Gómez criteria9, presence of comorbidities or immunosuppressive conditions, and lack of informed consent. |
| Rahman-2017 | HIV negative primary VL patients between 5 and 60 years screened with positive rK39 rapid immunochromatographic tests (InBios, Seattle, USA) and parasitologically confirmed via bone marrow or spleen aspirates (only at CBMC) were enrolled into the study after giving informed consent.  Because of the potential teratogenic effects of miltefosine, women of childbearing age who were not using an assured method of contraception for the duration of treatment and three months afterwards were excluded, unless they agreed to receive an injection of medroxyprogesterone acetate (DepoProvera, Pfizer, NY, USA).One injection (which is effective for 3 months) was needed to ensure adequate coverage, taking into account miltefosine's long half-life of approximately 7 days. Single women of childbearing age were randomized to receive either AmBisome alone or AmBisome + paromomycin.  Other exclusion criteria were: known hepatitis B, hepatitis C, or HIV infection, Hb concentrations less than 5 g/dl, platelet count of less than 40,000/mm3 (at CBMC only), a prothrombin time of more than 5 seconds longer than the control (at CBMC only), severe malnutrition [for adults (> 18 years) defined as BMI <14; for children (< 18 years) defined as BMI for age z score < -3 in children measuring > 121.5cm; and weight for height less than 60% in children measuring <121.5 cm], known alcohol or drug abuse, use of any investigational (unlicensed) drug within the last 3 months, and severe concurrent illnesses (TB, malaria) or chronic conditions (diabetes, hypertension). Pregnant and breast-feeding women, and patients with known hypersensitivity to the study drugs were also excluded |
| Romero-2017 | Exclusion criteria were pregnancy; HIV infection; underlying chronic or acute disease which would preclude evaluation of the participant's response to study medication (e.g. diabetes, cardiac, renal, or hepatic impairment, schistosomiasis, malaria, tuberculosis); co-morbidities that may cause alterations of the immune system; any concomitant use of medication that may interfere with the therapeutic response or cause detrimental pharmacological interactions; previous treatment with any anti-leishmanial drugs in the past 6 months; drug abuse; previous history of hypersensitivity reaction to tested interventions; any condition that may hinder compliance with the planned scheduled visits; relapse cases; clinical signs of severe VL disease, such as generalized edema, jaundice, toxemic signs, and severe malnutrition; serum creatinine and bilirubin above the upper normal limit (UNL); prolonged prothrombin time with international normalized ratio (INR) > 2.0; or a platelet count < 20,000/mm. |
| Alborzi-2017 | Inclusion criteria were signs of infection (fever, splenomegaly, etc.), positive serology IFA titer 1/128 or rK39 strip test, and/or positive bone marrow microscopy for Leishman bodies. Exclusion criteria were presence of clinically obvious jaundice, disseminated intravascular coagulation (DIC), and/or shock |
| Pandey-2017 | Participants of either sex, aged less than 15 years, and having clinical signs and symptoms consistent with kala-azar and positive rK39 test (Inbios) were included. The diagnosis was confirmed by parasitological analysis of splenic or bone marrow aspiration. Only parasitologically confirmed cases were included in the study. Fourteen patients had a past history of VL for which they had been treated with different antileishmanial drugs (Table 1)  Exclusion criteria were hemoglobin (Hb) < 5 g/100 mL; white blood cells count < 1,000/mm3; thrombocyte count < 50,000/mm3; alanine transaminase, aspartate transaminase, and alkaline phosphatase > 2.5 times upper limit of normal range; bilirubin ³ 2 times upper limit of normal, serum creatinine or blood urea nitrogen ³ 1.5 times upper limit of normal. Patients seropositive for HIV, hepatitis B and C, hypersensitivity to amphotericin B or inactive ingredients of the amphotericin B formulation were also excluded. |
| Kimutai-2017 | Only consenting patients treated with a SSG-PM combination were enrolled. Patients did not receive the combination if they had severe renal, cardiac or other systemic disease based on clinician judgment, or if they were known to be taking an aminoglycoside or were hypersensitive to either SSG or PM. As per MSF clinical protocol, HIV coinfected patients and patients above 50 years of age received AmBisome rather than SSG-PM in MSF sites. In other sites, those patients who received SSG-PM were thus included in the PV programme. |
| Mbui-2018 | Each recruited patient fulfilled the following inclusion criteria: (i) clinical signs and symptoms of VL and confirmatory parasitological microscopic diagnosis, (ii) aged ≥4 years and ≤12 years, (iii) able to comply with the study protocol, (iv) written informed consent signed by parents(s) or legal guardian, and children`s assent (>8-12 years, in Uganda only), (v) body weight of <30 kg.  None suffered severe malnutrition or any serious underlying disease or concomitant severe infection. None had received anti-leishmanial drugs within the previous 6 months.  From Supplementry file: They each had none of the following exclusion criteria: (i) relapsed VL, (ii) received any anti-leishmanial drugs within the previous six months, (iii) severe malnutrition (= a z score of <-3 on weight-for-height WHO reference curves, if aged <5 years or a z score of <-3 on BMI-for-age WHO reference curves, if aged 5-12 years), (iv) positive HIV diagnosis, (v) previous history of hypersensitivity to miltefosine, (vi) concomitant severe infection or any other serious underlying disease, (vii) any condition associated with splenomegaly, such as schistosomiasis, (viii) had reached menarche (female patients), (ix) haemoglobin concentrations of <5 g/dL, (x) white blood cell count of <1x103/mL blood, (xi) platelet count of <40,000/mL blood, (xii) abnormal liver function (= blood alanine-transferase and aspartate transferase levels >3-times above upper limit of normal range), (xiii) blood bilirubin levels >1.5-times greater than upper limit of normal range, (xiv) serum creatinine levels above upper limit of normal range, (xv) clinical signs of severe VL (jaundice and bleeding), (xvi) unable to comply with study protocol. |
| Goyal-2018 | All patients meeting a case definition of VL defined as fever for more than 2 weeks, splenomegaly, and confirmed with a positive rK-39 rapid diagnostic test (InBios, USA) were included in the study. Relapse cases with a confirmatory parasitological diagnosis were also eligible.  Patients with concurrent PKDL, HIV and those reporting a history of hypersensitivity to the investigational drugs were excluded. Patients with haemoglobin <4 g/dl, serious concomitant infection (e.g. severe pneumonia), complicated severe malnutrition, TB/VL co-infection, or children <2 years of age were referred to the MSF VL treatment unit within Hajipur district hospital or RMRIMS for further specialist management. These patients were treated with SDA as per physician decision and included in the study. |
| Sundar-2019 | Patients of all ages and of both genders, having signs and symptoms suggestive of VL, that is, fever with chills and rigor for 2 weeks or more, not responding to antimalarial drugs, and splenomegaly were included in the study. An rK39 antigen–based immunochromatographic serological rapid test was performed. Patients with typical signs and symptoms with characteristic laboratory features (pancytopenia) and a positive strip test were included in this study. Patients with serious illnesses or concurrent infections, such as tuberculosis or bacterial pneumonia, HIV, and Hepatitis B/C, and known allergy to AmB were excluded |
| Diro-2019 | • Confirmed HIV positive test (2 rapid diagnostics tests (RDTs) followed by a confirmatory ELISA test). • Diagnosis of VL (first episode or relapse) confirmed by bone marrow or spleen aspirate.* • Male and female age: 18-60 years. • Written informed consent from the patient.  (NOTE: information in supp. file)  The presence of any of the following will exclude a patient from study enrolment: • Women of child-bearing potential (defined as women who have achieved menarche) who are not using an assured method of contraception or are unwilling to use an assured method of contraception for the duration of treatment and four months after.*** • Pregnant women or breast-feeding mothers. • Patients with grade 2 or 3 post kala-azar dermal leishmaniasis (PKDL) lesions. • Clinical or biological evidence of severe cardiac, renal or hepatic impairment. • Known hypersensitivity to AmBisome® and/or miltefosine. • Patients receiving allopurinol treatment |
| Sinha-2019 | We included children, 2-11 years of age of both male and female sex, and had VLsuspected on the basis of clinical presentation (fever, splenomegaly determined by palpation, anemia, and cytopenia) and conrmed by the demonstration of amastigotes in splenic aspirate specimens  Exclusion criteria were values for the formed elements of the blood suggesting a premorbid state and severe disease (platelet count, <50,000 platelets/μL; leukocyte count, <1000 leukocytes/μL; hemoglobin concentration, <6 g/dL; serum aspartate aminotransferase (ASAT) or alanine aminotransferase levels of >3 times the upper limit of normal; bilirubin level of more than twice the upper limit of normal; and serum creatinine or blood urea nitrogen levels of >1.5 times the upper limit of normal), documentation of other serious coincidental medical illnesses, HIV seropositivity, and, for patients capable of reproduction, pregnancy. |
| Goswami-2020 | Patients of both genders aged between 5 and 65 years with a corroborative clinical history (patients from endemic areas with a prolonged fever not responding to antimalarials antibiotics), physical signs (anemia, splenomegaly, and hepatomegaly), and presence of parasites confirmed by examination of Giemsa-stained slides of splenic or bone marrow aspirates were enrolled into the study.  The following groups of patients were excluded: HIV-positive individuals, infants and children with body weight < 10 Kgs, patients with severe concurrent illnesses, patients who received any antileishmanial drugs or antifungal drugs in the previous 45 days, pregnant patients, and those with withdrawal of contraceptive measures. We also excluded patients with known hypersensitivity to the study drugs and those with diabetes, hypertension, tuberculosis, and heart, liver, or kidney disease |
| Ekram-2021 | After taking consent either from the patients or parents (or guardians) of the child, all consecutive adult and paediatric cases (age range 3-65 years) suspected of VL were approached for inclusion into the study. During final inclusion, parasites found on microscopy of a splenic aspirate smear or bone marrow was considered the prime selection criteria  Included: patients who were seropositive for HIV, and had a serious concurrent infection (e.g. TB or bacterial pneumonia) (Zahra et al. 2018), a serum creatinine level of [1.5 times the upper limit of the normal range, or have a history of allergy or hypersensitivity to amphotericin B. Moreover, anti-leishmanial or unlicensed investigational treatments within 180 days, pregnant women and patient and/or attendant refusing to give consent to take part in the study were also excluded |
| Haidar-2001 | The inclusion criteria were: all the confirmed cases of visceral leishmaniasis up to 12 years of age with an agreement from the parents or relatives for follow-up. History of taking antimalarial drugs was positive in 47%. |

**Table 2: Assessment of nutritional status of the patients**

| **Author year** | **Criteria used for assessment of nutritional status** |
| --- | --- |
| Ritmeijer-2006 | Defined based on BMI |
| Hailu-2010 | Showed severe protein or caloric malnutrition (Kwashiorkor or marasmus); *Children: Weight for age defined as severely underweight if ,60%, underweight if between 60% and 80%, normal if .80%. Adults: BMI defined as severely underweight if ,16, underweight if between 16.0 and 18.4, normal if between 18.5 and 24.9 |
| Musa-2010 | Severe protein or caloric malnutrition (Kwashiorkor or marasmus);  Based on Weight for Height (WHO child growth standards) if age ,6 years) and BMI for Age if age 6–19 years; defined as severely underweight if z-score,23SD; underweight if 23SD#z-score ,22SD; normal if 22SD#z-score,+1SD; and BMI if age .19 years: defined as severely underweight if ,16, underweight: 16.0–18.4, normal: 18.5–24.9 |
| Dietze-2001 | severe protein and/ or caloric malnutrition (kwashiorkor or marasmus) |
| Mueller-2008 | Categorization of nutritional status was based either on weight-for-height, for girls who were ,137 cm tall and boys who were ,145 cm tall, or on body mass indexes (BMI). Weights-for-height that were ,70%, 70%–79%, 80%–89% and .89% of ‘normal’ (Anon. 1977) or BMI of ,16, 16–17, .17–18, and .18 were taken as indications of severe, moderate, mild, and no malnutrition, respectively. The statistical analyses were performed using the STATA software package (StataCorp, College Station, TX). |
| Khalil-2014 | Classified using weight for height and BMI for age in those aged #19 years and BMI in those aged .19: normal if 22SD# weight for height or BMI for age, +1SD or 18.5#BMI,25.0; underweight if 23SD# weight for height or BMI for age ,22SD or 16.0#BMI,18.5; severely underweight if weight for height or BMI for age ,23SD or BMI,16.0. |
| Musa-2012 | Nutritional status was classified as normal, underweight, or severely underweight according to WHO Child Growth Standards in those ,19 years and body mass index (BMI) in those >20 years [18]. |
| Veeken-2000 | Based on BMI and WHZ |
| Sinha-2010 | Patient nutritional status, as measured by MUAC, improved significantly from study enrolment to follow-up periods. No patient had a MUAC measurement ≤ 135 mm at 6 months follow-up. The mean BMI at enrolment among adults aged ≥ 17 years was 18.3 (SD ± 2.5). By 6 months, their mean BMI was approximately 20.4, an increase from enrolment of 2.1 (SD ± 1.8; P < 0.0001). Children and adolescents ≤ 16 years at enrolment were, on average, in the 19th BMI percentile. Their BMI increased by 19 BMI percentile points at 6 month follow-up ( P < 0.0001). |
| Mueller-2007 | WHZ and BMI |
| Mbui-2018 | Based on WHO standardized nutritional status. Children with severe malnutrition were excluded (z‐score <−3). For children aged <5 years, weight for height was used: children were considered underweight when their z‐score <−2 and overweight when their z‐score >2. For children aged 5–12 years, body mass index for age was used: children were considered underweight when their z‐score <−2 and overweight when their z‐score >1. |
| Borges-2017 | Severe malnutrition according to Gómez criteria, presence of comorbidities or immunosuppressive conditions |
| Wasunna-2016 | Severe protein and/or caloric malnutrition defined as kwashiorkor or marasmus in children and BMI <15 in adults.  *BMI in kg/m2; **nutritional status categorization derived by post-hoc determination of WHO standardized value, as opposed to the BMI threshold in the severely underweight exclusion criterion. |
| Rahman-2017 | Severe malnutrition [for adults (> 18 years) defined as BMI <14; for children (< 18 years) defined as BMI for age z score < -3 in children measuring > 121.5cm; and weight for height less than 60% in children measuring <121.5 cm] |
| Goyal-2018 | Anthropometric indicators appropriate for patient age were calculated using the latest World Health Organization (WHO) Multicentre Growth Reference [17]. Severe wasting was defined based on WHO criteria (weight for height Z-score <-3 for children <5 years; BMI-for-age Z-score <-3 for those 5–19 years; and BMI <16.0 for adults). |
| Ekram-2021 | Gross underweight was measured by BMI |

**Table 3: Description of severe disease or critical illness**

| **Author year** | **Extract** |
| --- | --- |
| Thakur-2000a | Definitions for **critical illness** from leishmaniasis included the spleen reaching to the pelvic crest, haemoglobin concentration < 50 g/l, white cell count < 2 × 109/l,3 platelet count < 80 × 109 /l,3 aspartate aminotransferase concentration > 4 × upper limit of normal, serum albumin concentration < 20 g/l, and urine urea or creatinine concentration > 2 × upper limit of normal |
| Thakur-2000b | Definitions for **critical illness** from leishmaniasis included the spleen reaching to the pelvic crest, haemoglobin concentration < 50 g/l, white cell count < 2 × 109 /l3, platelet count < 80 × 109 /l3, aspartate aminotransferase concentration > 4 × upper limit of normal, serum albumin concentration < 20 g/l, and urine urea or creatinine concentration > 2 × upper limit of normal. |
| Moore-2001 | It may be noted that patients were included even if they had **severe kala-azar**, or comorbidity such as anaemia or cachexia. Pregnancy, breastfeeding, advanced age and infancy were not reasons for exclusion. Patients with a past history of kala-azar or who had received any antimonial in the past were excluded. Patients who were clinically suffering from kala-azar but had both a negative DAT test and a negative splenic aspirate were also excluded. |
| Bhattacharya-2004 | Exclusion criteria were values for the formed elements of the blood suggesting a premorbid state and **severe disease** (platelet count, <50,000 platelets/mL; leukocyte count, <1000 leukocytes/mL; haemoglobin concentration, <6 g/dL; serum aspartate aminotransferase (ASAT) or alanine aminotransferase levels of >3 times the upper limit of normal; bilirubin level of more than twice the upper limit of normal; and serum creatinine or blood urea nitrogen levels of >1.5 times the upper limit of normal) |
| Wasunna-2005 | Females that were pregnant or lactating were excluded. Additional exclusion criteria included known hypersensitivity to sitamaquine, use of an anti-leishmanial agent within 10 days or of an investigational compound within 30 days (or 5 half-lives) of the start of the study, subjects with marked deviations in serum chemistry or abnormal hematologic markers outside those expected to be caused by VL, patients with serious underlying disease, including human immunodeficiency virus (HIV) infection, malnutrition, or **severe kala-azar,** glucose-6-phosphate dehydrogenase deficiency, or previous inclusion in this study. |
| Jha-2005 | Exclusion criteria included known hypersensitivity to sitamaquine, receipt of an antileishmanial agent within 10 days or of an investigational compound within 30 days (or 5 half-lives) of study start, sub- jects with marked deviations in serum chemistry, patients with serious underlying disease, including HIV infection, mal- nutrition or **severe kala-azar,** pregnancy or lactation, G6PD deficiency, or previous inclusion in this study. |
| Sundar-2011a | Patients of age 2–65y with **mild-moderate kala-azar**; lack of concomitant diseases such as malaria, HIV, and tuberculosis; lack of pregnancy or lactation; no previous treatment with antimony or paromomycin unless the treatment terminated two months prior to the study and the patient was worsening |
| Musa-2012 | Briefly, patients aged 4–60 years with parasitologically confirmed VL were included, but patients with **very severe VL** or those with contraindications were excluded. Patients were excluded if they: (1) had taken any antileishmanial drug in the preceding 6 months; (2) showed severe protein or caloric malnutrition (Kwashiorkor or marasmus); (3) had previous hypersensitivity reaction to SSG or aminoglycosides; (4) had a concomitant severe infection (except HIV) or any other serious underlying (cardiac, renal, hepatic) disease; (5) had conditions associated with splenomegaly, such as schistosomiasis; (6) had a history of cardiac arrhythmia or an abnormal electrocardiogram (ECG); (7) were pregnant or lactating; (8) had any relevant outliers of safety laboratory parameters (haemoglobin <5 g/dL, white blood cells <1�103/mm3, platelets <40,000/mm3, liver function test values >3 times higher than upper limit of normal, serum creatinine values above upper limit of normal); or (9) had clinical hearing loss. |
| Khalil-2014 | Patients with age of at least 4 years, confirmed HIV-negative, parasitologically-confirmed **non-severe VL,** were enrolled in three centres  Exclusion criteria were signs/symptoms of **severe VL** (patients who were very weak, unable to walk, bleeding, jaundiced, suffering from sepsis and other concomitant infections/illnesses); anti-leishmanial or unlicensed investigational treatments within six months; underlying chronic disease such as severe cardiac, pulmonary, renal, or hepatic impairment; serum creatinine outside the normal range; liver function tests more than 3 times the normal range; platelet count less than 40,000/mm3; known alcohol abuse; pregnancy or lactation; concomitant acute drug usage for malaria and bacterial infection; pneumonia within last 7 days; known hypersensitivity to AmBisome or amphotericin B; any other condition which may invalidate the trial |
| Romero-2017 | Clinical signs of **severe VL disease**, such as generalized edema, jaundice, toxemic signs, and severe malnutrition; serum creatinine and bilirubin above the upper normal limit (UNL); prolonged prothrombin time with international normalized ratio (INR) > 2.0; or a platelet count < 20,000/mm. |
| Mbui-2018 | They each had none of the following exclusion criteria: (i) relapsed VL, (ii) received any anti-leishmanial drugs within the previous six months, (iii) severe malnutrition (= a z score of <-3 on weight-for-height WHO reference curves, if aged <5 years or a z score of <-3 on BMI-for-age WHO reference curves, if aged 5-12 years), (iv) positive HIV diagnosis, (v) previous history of hypersensitivity to miltefosine, (vi) concomitant severe infection or any other serious underlying disease, (vii) any condition associated with splenomegaly, such as schistosomiasis, (viii) had reached menarche (female patients), (ix) haemoglobin concentrations of <5 g/dL, (x) white blood cell count of <1x103/mL blood, (xi) platelet count of <40,000/mL blood, (xii) abnormal liver function (= blood alanine-transferase and aspartate transferase levels >3-times above upper limit of normal range), (xiii) blood bilirubin levels >1.5-times greater than upper limit of normal range, (xiv) serum creatinine levels above upper limit of normal range, (xv) **clinical signs of severe VL** (jaundice and bleeding), (xvi) unable to comply with study protocol. |
| Sinha-2019 | Exclusion criteria were values for the formed elements of the blood suggesting a premorbid state and **severe disease** (platelet count, <50,000 platelets/μL; leukocyte count, <1000 leukocytes/μL; hemoglobin concentration, <6 g/dL; serum aspartate aminotransferase (ASAT) or alanine aminotransferase levels of >3 times the upper limit of normal; bilirubin level of more than twice the upper limit of normal; and serum creatinine or blood urea nitrogen levels of >1.5 times the upper limit of normal) |

**Table 4: Case definitions and case confirmation approach**

| **Author year** | **case definition method** | **compatible signs and symptoms** | **Cas confirmation method** |
| --- | --- | --- | --- |
| Gaeta-2000 | Compatible clinical diagnosis | fever + splenomegaly + pancytopenia | Parasitological |
| Sundar-2000c | Compatible clinical diagnosis | fever + splenomegaly + weight loss + weakness | Parasitological |
| Thakur-2000a | Compatible clinical diagnosis | fever + splenomegaly/hepatosplenomegaly + weight loss/loss of appetite | Parasitological |
| Sundar-2000b | Compatible clinical diagnosis | fever + weight loss + hepatosplenomegaly + pancytopenia | Serological and/or parasitological |
| Sundar-2000a | Compatible clinical diagnosis | fever + splenomegaly/hepatosplenomegaly + weight loss/loss of appetite | Serological and/or parasitological |
| Thakur-2000b | Compatible clinical diagnosis | Unclear | Serological and/or parasitological |
| Veeken-2000 | Compatible clinical diagnosis | fever + splenomegaly/wasting | Serological and/or parasitological |
| Villanueva-2000 | Not presented | Unclear | Parasitological |
| Haidar-2001 | Compatible clinical diagnosis | Unclear | Serological and/or parasitological |
| Thakur-2001a | Compatible clinical diagnosis | fever + splenomegaly | Parasitological |
| Moore-2001 | Compatible clinical diagnosis | fever + splenomegaly/wasting | Serological and/or parasitological |
| Ritmeijer-2001 | Compatible clinical diagnosis | fever + splenomegaly/wasting | Serological and/or parasitological |
| Thakur-2001b | Not presented | Unclear | Parasitological |
| Dietze-2001 | Compatible clinical diagnosis | Unclear | Serological and/or parasitological |
| Sundar-2001 | Compatible clinical diagnosis | fever + splenomegaly/hepatosplenomegaly + weight loss/loss of appetite | Serological and/or parasitological |
| Das-2001 | Not presented | Unclear | Parasitological |
| Sundar-2002 | Compatible clinical diagnosis | fever + splenomegaly + cytopenia | Parasitological |
| Sundar-2002a | Compatible clinical diagnosis | Unclear | Serological and/or parasitological |
| Laguna-2003 | Compatible clinical diagnosis | Unclear | Parasitological |
| Sundar-2003b | Compatible clinical diagnosis | fever + chills + rigor + splenomegaly | Serological and/or parasitological |
| Figueras Nadal-2003 | Compatible clinical diagnosis | Unclear | Serological and/or parasitological |
| Sundar-2003 | Compatible clinical diagnosis | Unclear | Serological and/or parasitological |
| Syriopoulou-2003 | Compatible clinical diagnosis | fever + splenomegaly + anaemia + leukopenia/thrombocytopenia | Serological and/or parasitological |
| Rijal-2003 | Compatible clinical diagnosis | fever + splenomegaly | Serological and/or parasitological |
| Thakur-2004a | Not presented | Unclear | Parasitological |
| Sundar-2004 | Compatible clinical diagnosis | fever + splenomegaly/hepatosplenomegaly + weight loss/loss of appetite | Parasitological |
| Thakur-2004b | Not presented | Unclear | Parasitological |
| Bhattacharya-2004 | Compatible clinical diagnosis | fever + splenomegaly + anaemia + cytopenia | Serological and/or parasitological |
| Wasunna-2005 | Compatible clinical diagnosis | Unclear | Parasitological |
| Jha-2005 | Compatible clinical diagnosis | Unclear | Parasitological |
| Das-2005 | Compatible clinical diagnosis | fever + splenomegaly | Serological and/or parasitological |
| Ritmeijer-2006 | Compatible clinical diagnosis | fever + wasting + splenomegaly/lymphadenopathy | Serological and/or parasitological |
| Sundar-2006 | Compatible clinical diagnosis | fever + splenomegaly | Parasitological |
| Singh-2006 | Compatible clinical diagnosis | fever + splenomegaly | Serological and/or parasitological |
| Singh-2006 | Compatible clinical diagnosis | fever + splenomegaly | Serological and/or parasitological |
| Sundar-2007b | Compatible clinical diagnosis | fever + splenomegaly/hepatosplenomegaly + weight loss/loss of appetite | Parasitological |
| Sundar-2007a | Compatible clinical diagnosis | Unclear | Serological and/or parasitological |
| Bhattacharya-2007 | Compatible clinical diagnosis | fever or splenomegaly or cytopenia | Serological and/or parasitological |
| Mueller-2007 | Compatible clinical diagnosis | fever + weight loss + splenomegaly/lymphadenopathy + hepatomegaly + epistaxis + anaemia | Serological and/or parasitological |
| Sundar-2008b | Compatible clinical diagnosis | fever + splenomegaly/hepatosplenomegaly + weight loss/loss of appetite | Parasitological |
| Sundar-2008a | Compatible clinical diagnosis | fever + splenomegaly/hepatosplenomegaly + weight loss/loss of appetite | Parasitological |
| Thakur-2008 | Not presented | Unclear | Parasitological |
| Mueller-2008 | Compatible clinical diagnosis | Unclear | Serological and/or parasitological |
| Sundar-2009a | Compatible clinical diagnosis | fever + splenomegaly/hepatosplenomegaly + weight loss/loss of appetite | Parasitological |
| Das-2009 | Compatible clinical diagnosis | fever + rigor + hepatosplenomegaly + anorexia + weight loss + anaemia | Parasitological |
| Sundar-2009b | Compatible clinical diagnosis | fever + splenomegaly/hepatosplenomegaly + weight loss/loss of appetite | Serological and/or parasitological |
| Adam-2009 | Compatible clinical diagnosis | fever + weight loss + anaemia + hepatosplenomegaly | Serological and/or parasitological |
| Shahian-2009 | Compatible clinical diagnosis | fever + hepatomegaly/splenomegaly | Serological and/or parasitological |
| Hailu-2010 | Compatible clinical diagnosis | fever + splenomegaly | Serological and/or parasitological |
| Thakur-2010 | Compatible clinical diagnosis | fever + shivering + hepatosplenomgaly + lukopenia | serological + parasitological |
| Musa-2010 | Compatible clinical diagnosis | Unclear | Serological and/or parasitological |
| Sundar-2010 | Compatible clinical diagnosis | fever + splenomegaly/hepatosplenomegaly + weight loss/loss of appetite | Serological and/or parasitological |
| Mondal-2010 | Compatible clinical diagnosis | fever + hepatomegaly/splenomegaly | serological + parasitological |
| Rijal-2010 | Compatible clinical diagnosis | fever + splenomegaly | Serological and/or parasitological |
| Singh-2010 | Compatible clinical diagnosis | Unclear | Serological and/or parasitological |
| Sinha-2010 | Compatible clinical diagnosis | fever + splenomegaly + weight loss + anaemia | Serological and/or parasitological |
| Sundar-2011b | Compatible clinical diagnosis | fever + splenomegaly | Serological and/or parasitological |
| Sundar-2011c | Compatible clinical diagnosis | fever + splenomegaly/hepatosplenomegaly + weight loss/loss of appetite | Parasitological |
| Sundar-2011a | Not presented | Unclear | Parasitological |
| Rahman-2011 | Compatible clinical diagnosis | fever + splenomegaly/hepatosplenomegaly + weight loss/loss of appetite | Serological and/or parasitological |
| Sinha-2011 | Compatible clinical diagnosis | Unclear | Serological and/or parasitological |
| Sudarshan-2011 | Not presented | Unclear | Parasitological |
| Sundar-2012 | Compatible clinical diagnosis | fever + chills + rigor + splenomegaly | Serological and/or parasitological |
| Musa-2012 | Compatible clinical diagnosis | fever + splenomegaly | Serological and/or parasitological |
| Patra-2012 | Compatible clinical diagnosis | fever + hepatosplenomegaly + anaemia + lymphadenopathy | serological + parasitological |
| Rijal-2013 | Compatible clinical diagnosis | fever + splenomegaly | Serological and/or parasitological |
| Khalil-2014 | Compatible clinical diagnosis | fever or splenomegaly or cytopenia | Serological and/or parasitological |
| Ostyn-2014 | Not presented | Unclear | Serological and/or parasitological |
| Cota-2014 | Compatible clinical diagnosis | fever or splenomegaly or cytopenia | Serological and/or parasitological |
| Sundar-2014 | Compatible clinical diagnosis | fever + splenomegaly | serological + parasitological |
| Mondal-2014 | Compatible clinical diagnosis | fever + splenomegaly | Serological and/or parasitological |
| Jamil-2015 | Compatible clinical diagnosis | fever + splenomegaly/hepatosplenomegaly + weight loss/loss of appetite | Serological and/or parasitological |
| Sundar-2015 | Not presented | Unclear | Parasitological |
| Goswami-2016 | Compatible clinical diagnosis | fever + splenomegaly | serological + parasitological |
| Wasunna-2016 | Not presented | Unclear | Parasitological |
| Pandey-2016 | Compatible clinical diagnosis | fever + splenomegaly | Serological and/or parasitological |
| Borges-2017 | Compatible clinical diagnosis | fever + splenomegaly | Serological and/or parasitological |
| Rahman-2017 | Not presented | Unclear | serological + parasitological |
| Romero-2017 | Compatible clinical diagnosis | fever + hepatomegaly/splenomegaly | Serological and/or parasitological |
| Alborzi-2017 | Compatible clinical diagnosis | fever + splenomegaly | Serological and/or parasitological |
| Pandey-2017 | Compatible clinical diagnosis | Unclear | serological + parasitological |
| Kimutai-2017 | Compatible clinical diagnosis | Unclear | Serological and/or parasitological |
| Mbui-2018 | Compatible clinical diagnosis | Unclear | Serological and/or parasitological |
| Goyal-2018 | Compatible clinical diagnosis | fever + splenomegaly | Serological and/or parasitological |
| Sundar-2019 | Compatible clinical diagnosis | fever + chills + rigor + splenomegaly | Serological and/or parasitological |
| Diro-2019 | Compatible clinical diagnosis | fever + splenomegaly/hepatosplenomegaly + weight loss/loss of appetite | Serological and/or parasitological |
| Sinha-2019 | Compatible clinical diagnosis | fever + splenomegaly + anaemia + cytopenia | Serological and/or parasitological |
| Goswami-2020 | Compatible clinical diagnosis | fever + splenomegaly + hepatomegaly + anaemia | Serological and/or parasitological |
| Ekram-2021 | Compatible clinical diagnosis | Unclear | Serological and/or parasitological |

**Table 5: Sample size estimation details**

| **Author year** | **Details of the sample size estimation** |
| --- | --- |
| Veeken-2000 | We calculated that, at a power of 90% and a significance level of P= 0.05 (two-tailed), to detect a difference of 10% in cure, death, or relapse rate between the two groups, a sample size of 207 patients in each arm was required. |
| Moore-2001 | Makuch & Simon's formula (3, 4) can be used to calculate sample sizes for comparative studies if the objective is to show that two treatments are equally effective. For this purpose, the difference in outcome that would lead one ofthe treatments to be discarded as inferior needs to be specified. For kala-azar treatment we judged this to be a 20% difference in cure rate. Since PSM has repeatedly been found to have a cure rate of about 95% for kala-azar in Africa, we considered that a 75% cure rate for generic sodium stibogluconate would be clinically unacceptable. Taking a = 0.05 and b = 0.1, we required 25 patients per treatment group. We also tested a second set of assumptions suitable for an unpaired prospective study with a dichotomous outcome (cure versus failure). Assuming the drugs differ by 20% in their cure rate, 95% in one group versus 75% in the other (a = 0.05; b = 0.2), we required 49 patients per treatment group. |
| Ritmeijer-2001 | A sample size of 91 patients in each arm was used, with a 90% power to detect at a significance level of P = 0.05 (2-tailed), a difference of 20% in cure, death, or relapse rate between the 2 group. |
| Das-2001 | Assuming a difference of 15% in the efficacy rate between two regimen groups and at level of significance p=0.05 and power of the test at 90%, a sample size of 158 patients unresponsive to antimony were allocated randomly into two groups, using random number table. |
| Sundar-2002 | The considerations for the sample size and the confirmatory analysis were determined on the basis of the restricted maximum-likelihood estimation of the variance of the test statistic, 15 with a one-sided alpha of 0.025, a power of 0.80, a margin of noninferiority of 15 percent, assumed cure rates for miltefosine of 88 to 92 percent and for amphotericin of 94 to 98 percent, and a ratio of 3:1 for the random assignment of patients to miltefosine and amphotericin. The number of patients required overall was 400. |
| Sundar-2002a | The sample size calculation was based on a complete cure rate hypothesis of 90% for the highest dose and 50% for the lowest dose. To detect a difference of 40% in cure rates with a power of 80% and a significance level of 5% in a 2-tailed approach, 25 patients per treatment arm were required |
| Sundar-2003b | Sample-size calculation was based on the definitive-cure rate of 90%, with a one-sided lower-bound CI of 0.84, a power of 80%, and a significance level of 5%; for this, a minimum of 200 patients were required |
| Thakur-2004a | The target sample size, set assuming there would be no drop-outs or spontaneous cures and that the inter-drug difference in the frequencies of cure would be 20% (Dowd, 1979), was 120 patients (i.e. 60/treatment arm) |
| Wasunna-2005 | Assuming a true response rate of 85%, this calculation showed that 61 patients would be required to detect within 9% of the true response rate at a confidence level of 95%, assuming that the binomial distribution can be approximated by a normal curve. It was subsequently decided to present exact 95% confidence inter- vals for the primary efficacy outcome calculated using the Clopper Pearson method. With 61 patients and assuming a true response rate of 85%, this method would detect within −11% to +8% of the true response rate at a two-sided confidence level of 95% |
| Sundar-2007b | The proportion of patients with a final cure determined the end point of this trial, which was designed to assess noninferiority of the experimental regimens. With a 1-sided a of 0.05, a power of 90%, and a margin of noninferiority of 5%, samples sizes were determined assuming cure rates of 91%-95% for 15 daily infusions of 0.75 or 1 mg/kg of amphotericin B and of95%-99% for 15 alternate- day infusions at the same doses and a ratio of 2:1 for random assignment to daily and alternate-day infusions. |
| Sundar-2007a | Assuming a 99% cure rate for amphotericin, 666 patients were needed in a 3:1 ratio to support a one-sided, non- inferiority analysis without stratification and with 80% power to detect a type I error rate of 5%. |
| Sundar-2008b | The study was designed to have a 5% type 1 error and 95% power, considering a failure rate of and <10% to indicate adequate efficacy (the minimum detectable failure rate at the ß = 0.05 level) and a failure rate ≥ 25% to indicate insufficient efficacy. The boundaries of the test were calculated for H0 ( p= p0) and Ha (p < pa) with p0= 0.25 and pa=0.10. Based on simulations, we expected the sample path to cross the H0 rejection line with an average sample size of 40 patients and the H0 non-rejection line with an average sample size between 20 and 25 patients. When, after enrolling 45-46 patients per arm, all treatments appeared to be equally and highly effective, an additional 45 consecutive patients were enrolled and non-randomly assigned to a fifth regimen. |
| Thakur-2008 | We needed a sample size of at least 60 based on past experience and assuming a rate of drop out and spontaneous cure of zero and a difference in the rate of cure between the standard treatment and the new treatment of 20 per cent. |
| Sundar-2009a | Sample size was determined after discussion with regulatory authorities for this phase II study and Schedule Y (amended 2005) of Drug and Cosmetic act of India. |
| Das-2009 | We believed that the reported cure rate of AMB (based on the literature) would be 95% in the Amphotericin B group, while it would be about 75% in the Pentamidine group. Based on 90% power to detect a significant difference (P = 0.05, two-sided) 100 patients were required for the study. To compensate for a 10%, drop out, an optimum sample size for this clinical trial was 110 patients, which came to 55 patients in each group. |
| Sundar-2009b | Assuming a 95% cure rate after 21 days of treatment with paromomycin, 327 patients were needed in a 2:1 ratio to support a 1-sided noninferiority analysis without stratification and with 80% power to detect a type I error rate of 5%. |
| Hailu-2010 | At the start of the trial, it was estimated that 217 patients would be required per arm to detect a 10% difference in efficacy among HIV-uninfected patients at the 5% level, assuming 90% power, 95% efficacy in the reference (SSG) arm, and a 20% drop-out rate from the analysis (10% due to HIV co-infection and 10% due to loss to follow-up between end of treatment and 6-month assessment). |
| Thakur-2010 | The aim of this study was to reduce complications such as rigor and acute renal failure caused by treatment with only amphotericin B by simultaneous administration of 500mL of physiologic saline and 30 mL (60 meq/L) of KCl in adults and a correspondingly lower dose in children.  A sample size of 100 patients in each group was required for 90% power to detect a difference of 11% reduction in complications in the test regimen group compared with the control regimen group at 5% significance level. On the basis of our previous experience, the drop-out rate was assumed to be 15% at this center. When we adjusting for the compliance rate, the final sample size was estimated to be 115 patients in each group. |
| Sundar-2010 | Assuming a 99% cure rate for standard treatment with amphotericin B deoxycholate, we determined that we would need to enrol at least 400 patients in a 3:1 ratio to support a one-tailed noninferiority analysis without stratification and with a power of 90% to detect the probability of a type I error of 5%. |
| Sinha-2010 | The patient sample size was set at 250, which is sufficient, with an expected 10% loss to follow-up, to detect a treatment success rate of 95% with ± 4% precision. |
| Sundar-2011b | We assumed a definitive cure rate of 97% with the reference drug (amphotericin B) and a non-inferiority margin of 7% for the test groups. With a power of 90% and equal allocation ratio, the sample size per group was 148; on the assumption of a drop-out rate of 5%, 156 patients per group were needed, with a total sample size for four groups of 624 patients. A non-inferiority margin of 7% was chosen because 90% was thought to be the minimum acceptable rate of definitive cure; it was the rate attained by a single 5 mg/kg dose of liposomal amphotericin B in a previous trial. |
| Musa-2012 | The trial was designed to have90% power (b=0.1) to detect, at the 5% significance level (a=0.05), an absolute difference in efficacy of 15% between PM and SSG and 10% between SSG & PM and SSG regimens [16]. An 85% efficacy was assumed in the reference arm and adjusting for 10% HIV co-infection and 10% loss to follow- up at 6 months post end of treatment, it was estimated that 404 and 195 patients per arm were required for the respective comparisons. Being HIV-positive was not an exclusion criterion but the original protocol stated that there was to be a sufficient number of patients for a subgroup analysis excluding HIV patients (if deemed necessary). |
| Khalil-2014 | For the primary endpoint comparison, 120 patients per arm would provide 80% power to detect non-inferiority within a margin of 10%, assuming 95% cure in the reference arm, a one-sided alpha of 0?05 and 15% loss-to-follow-up. In interim analyses, 20 patients per arm would provide 90% power to detect a difference of at least 35% in parasite clearance rate at day 30, assuming 95% cure in the reference arm and a two-sided alpha of 0.05. With 40 patients per arm, there would be 90% power to detect a difference of at least 25% under the same assumptions. |
| Sundar-2014 | A total of 500 patients in a 3:1 ratio (375 in ABLE and 125 in LAmB) were planned to be enrolled assuming a dropout rate of 20% and non-inferiority margin fixed at 20.10. This was expected to provide an estimated difference in proportions of patients achieving definitive cure for ABLE vs. LAmB equals to zero, with at least 80% power for the non-inferiority test. |
| Mondal-2014 | A purposive sample size was determined, assuming that at least 85% of patients with visceral leishmaniasis who attended the hospital were eligible to be treated with single-dose liposomal amphotericin B, and aiming to obtain at least 95% cure rate at 6 months and with up to 3% of treated patients developing drug-related adverse events requiring referral to a tertiary hospital for management. |
| Goswami-2016 | A sample size of 75 patients per group was sufficient to detect a clinically important difference of 11% between groups in the definitive cure rate at 6 months using a two- sided Z-test with 80% power and 5% significance level. |
| Wasunna-2016 | The trial used a sequential design with a triangular continuation region [14]. The null hypothesis was that the proportion cured at day 28 (p) is less than or equal to a value p0 which we set to 75%. The alternative hypothesis is that p>p0. If the upper boundary is crossed during an interim analysis, then the null hypothesis is rejected and we conclude p>75%. Crossing the lower boundary at the time of an interim analysis implies that null hypothesis (proportion for which we chose a value of90%. The type I error rate and power of the study were pre-specified cured 75%) is not rejected and there is specified power to exclude a proportion cured as 5% and 95%, respectively (α = β = 0.05). Interim analyses were specified after every 15 patients in each arm. The maximum sample size per arm was 63. The study was designed and analyzed according to sequential methods, which have been developed to allow for discrete data analysis after a pre-specified number of patients are recruited. The triangular test is one such method and uses straight line stopping boundaries [13]. The continuation region is closed, which ensures a maximum sample size. A minimum sample size of30 per arm was imposed to allow for adequate PK assessment. The trial was non-comparative and the sequential analysis was applied to each arm independently, allowing them to potentially stop at different times. |
| Borges-2017 | The original sample size was determined on the basis of the primary outcome endpoint of efficacy. To calculate sample size, an ideal setting with unlimited number of patients was simulated and the formula suggested by Pocock was applied. The required sample size was 283 subjects in each group. As a pilot trial, this study was projected with 50 individuals in each arm to evaluate the protocol. |
| Rahman-2017 | The sample size was calculated assuming a treatment success of 97% in the reference arm (AmBisome) and a margin ofnon-inferiority of any tested treatment of 7%, leading to a minimally acceptable cure rate of 90% for each treatment. With a power of 90%, the sample size per group in the sample of married women, men and children would be 140. Based on the possible teratogenic effect of miltefosine and the assumption that 20% of the patients would be single women of child bearing potential, 26 extra patients were to be recruited among these women for both non-miltefosine groups. Assuming a drop-out rate of 10%, 154 patients were needed in both miltefosine groups and 183 in both non-miltefosine groups. The total sample size was calculated to be 674 patients. |
| Romero-2017 | As the trial objective was to compare three interventions with the standard treatment (MA), the sample size was determined to allow for detection, with 80% power (1- β) and 5% significance level (α = 0.05), of at least 8% difference in efficacy of each treatment arm in relation to the reference arm. Assuming a 90% efficacy in the MA reference arm, and adjusting for a 10% loss to follow-up and 10% to maintain the power of comparison between the patient subgroups with parasitological diagnostic confirmation and only a positive rk39 diagnostic test (as defined in the inclusion criteria), it was estimated 165 participants would be required per treatment arm, for a total sample size of 660 participants. After the AmphoB arm was stopped due to its higher toxicity, the trial's sample size was reviewed. Adjustment for loss to follow-up was reduced to 5%, in accordance with the observations from the trial up to that moment. No more adjustment was made for those patients diagnosed with only a positive rK39 test, due to the wide validation of these tests by TDR/WHO on three continents, which allows for the inclusion of patients with clinical signs and a positive immunochromatographic test [18]. A final total sample size of 426 was calculated, with 142 participants per treatment arm. |
| Kimutai-2017 | Given the emphasis on safety, the sample size required was based on the probability of detecting AEs with an expected incidence of 1/1000. With a cohort of 2000 patients, there would be an 86% probability of detecting at least one AE with an expected 1/1000 incidence and a 95% probability with a cohort of 3000 patients [20–22]. When the PV program reached a sample size of 3126, the Steering Committee held a final meeting to recommended termination of the PV data collection activities |
| Mbui-2018 | The minimal sample size was based on pharmacokinetic clinical trial simulations for the primary pharmacokinetic endpoint using the method of Dorlo and colleagues [18]. Including potential non-compliance, this provided a trial sample of 30 patients. For the primary pharmacokinetic endpoint, plasma miltefosine concentrations were measured in all patients receiving at least 1 miltefosine dose |
| Goyal-2018 | Since the objective of the study was to evaluate the effectiveness and safety of each new treatment modality, the sample size requirement was based around the precision with which effectiveness and safety could be estimated. Assuming a risk of failure of5% at 6-months follow-up, a sample size of225 patients per arm would allow for an effectiveness estimation with 3% precision. Since treatment modality allocation was planned to be different between sites, and the patient population might not be homogeneous (referral hospital vs PHC in different districts), an adjustment was applied using a conservative design effect of4 to account for between-centre variability. In this case, a failure risk of5% could be estimated at around 5% precision with 300 patients per arm. |
| Goswami-2020 | We assumed a definitive cure rate of 90.3% with the reference drug (miltefosine monotherapy) and a non-inferiority margin of 7% for the test groups.With a power of 90% and equal allocation ratio, the sample size per group was 53, with a total sample size of 106. A non-inferiority margin of 7% was chosen because 83% was thought to be the minimum acceptable rate of definitive cure as it was the rate attained by a phase IV trial of miltefosine for the treatment of Indian patients with VL |

**Table 5: Diagnostic methods used for enrolment and outcome assessment for relapse**

| **Author year** | **Diagnostic method used for**  **Disease confirmation** | **Tissue aspirates** | **QC of lab methods** | **Zymodeme analysis** | **Genotyping for  relapse** |
| --- | --- | --- | --- | --- | --- |
| Gaeta-2000 | Parasitological + culture | Bone marrow | No information | Not done | No |
| Sundar-2000c | Parasitological | Spleen | No information | Not done | No |
| Thakur-2000a | Parasitological | bone marrow and/or splenic aspirate | No information | Not done | No |
| Sundar-2000b | Parasitological | bone marrow and/or splenic aspirate | No information | Not done | No |
| Sundar-2000a | Parasitological | Spleen | No information |  | No |
| Thakur-2000b | Parasitological | bone marrow and/or splenic aspirate | No information |  | No |
| Veeken-2000 | Serological and/or parasitological | bone marrow or spleen or lymph node | Random samples checked |  | No |
| **Villanueva-2000** | Parasitological + culture | Combination | No information | Yes | No |
| Thakur-2001a | Parasitological | Spleen | No information | Not done | No |
| Moore-2001 | Serological and/or parasitological | Spleen | independent microscopist | Not done | No |
| Ritmeijer-2001 | Serological and/or parasitological | Spleen | independent microscopist | Not done | No |
| Thakur-2001b | Parasitological | bone marrow and/or splenic aspirate | No information |  | No |
| Dietze-2001 | Parasitological + culture | Spleen | No information | Not done | No |
| Sundar-2001 | Parasitological | Spleen | No information |  | No |
| Haidar-2001 | Parasitological | bone marrow and/or splenic aspirate | No information |  | No |
| Das-2001 | Parasitological | bone marrow and/or splenic aspirate | No information |  | No |
| Sundar-2002 | Parasitological | Spleen | No information | Not done | No |
| Sundar-2002a | Parasitological | bone marrow and/or splenic aspirate | No information | Not done | No |
| Laguna-2003 | Parasitological + culture | Combination | No information | Not done | Yes |
| Sundar-2003b | Parasitological | bone marrow and/or splenic aspirate | Single reader |  | No |
| Figueras Nadal-2003 | Serological and/or parasitological + culture | Bone marrow | No information | Not done | No |
| Sundar-2003 | Parasitological | Spleen | No information |  | No |
| Syriopoulou-2003 | Serological and/or Parasitological | Bone marrow | experienced technician used |  | No |
| Rijal-2003 | Parasitological | bone marrow and/or splenic aspirate | External checking of 10% of slides | Not done | No |
| Thakur-2004a | Parasitological | bone marrow and/or splenic aspirate | No information | Not done | No |
| Sundar-2004 | Parasitological | Spleen | No information | Not done | No |
| Thakur-2004b | Parasitological + culture | Bone marrow | No information | Not done | No |
| Bhattacharya-2004 | Parasitological | Spleen | No information |  | No |
| Wasunna-2005 | Parasitological | Spleen | No information | Not done | No |
| Jha-2005 | Parasitological | Spleen | No information | Not done | No |
| Das-2005 | Parasitological | bone marrow and/or splenic aspirate | No information |  | No |
| Ritmeijer-2006 | Serological and/or parasitological | Combination | No information | Not done | No |
| Sundar-2006 | Parasitological | Spleen | No information | Not done | No |
| Singh-2006 | Parasitological | Spleen | No information |  | No |
| Singh-2006 | Parasitological | Spleen | No information |  | No |
| Sundar-2007b | Parasitological | Spleen | No information | Not done | No |
| Sundar-2007a | Parasitological | bone marrow and/or splenic aspirate | No information | Not done | No |
| Bhattacharya-2007 | Parasitological | Spleen | No information | Not done | No |
| Mueller-2007 | Serological and/or parasitological | bone marrow or spleen or lymph node | No information |  | No |
| Sundar-2008b | Parasitological | Spleen | No information | Not done | No |
| Sundar-2008a | Parasitological | Spleen | No information | Not done | No |
| Thakur-2008 | Parasitological | bone marrow and/or splenic aspirate | No information | Not done | No |
| Mueller-2008 | Serological and/or parasitological | Spleen | No information |  | No |
| Sundar-2009a | Parasitological | bone marrow and/or splenic aspirate | No information | Not done | No |
| Das-2009 | Parasitological | bone marrow and/or splenic aspirate | No information | Not done | No |
| Sundar-2009b | Parasitological | Spleen | No information |  | No |
| Adam-2009 | Parasitological | Bone marrow | No information | No | No |
| Shahian-2009 | Serological | Unclear | No information | No | No |
| Hailu-2010 | Parasitological | bone marrow or spleen or lymph node | No information |  | No |
| Thakur-2010 | Serological and/or parasitological | bone marrow and/or splenic aspirate | No information |  | No |
| Musa-2010 | Parasitological | Bone marrow | Trained lab tech |  | No |
| Sundar-2010 | Parasitological | Spleen | Two readers | Not done | No |
| Mondal-2010 | Serological and/or parasitological | bone marrow and/or splenic aspirate | No information |  | No |
| Rijal-2010 | Parasitological | bone marrow and/or splenic aspirate | No information |  | No |
| Singh-2010 | Parasitological | Bone marrow | No information |  | No |
| Sinha-2010 | Serological and/or parasitological | Combination | No information |  | No |
| Sundar-2011b | Serological and/or parasitological | bone marrow and/or splenic aspirate | External checking of 10% of slides | Not done | No |
| Sundar-2011c | Parasitological | bone marrow and/or splenic aspirate | No information | Not done | No |
| Sundar-2011a | Parasitological | Spleen | No information | Not done | No |
| Rahman-2011 | Serological | Blood | No information |  | No |
| Sinha-2011 | Serological and/or parasitological | bone marrow or spleen or lymph node | No information |  | No |
| Sudarshan-2011 | Parasitological | Spleen | No information | No | PCR used on blood sample for parasite clearance but not for genotyping |
| Sundar-2012 | Parasitological | Spleen | Two readers | Not done | No |
| Musa-2012 | Parasitological | bone marrow or spleen or lymph node | No information |  | No |
| Patra-2012 | Serological and/or parasitological + culture | bone marrow and/or splenic aspirate | No information |  | No |
| Rijal-2013 | Parasitological + culture | bone marrow and/or splenic aspirate | No information |  | Yes |
| Khalil-2014 | Parasitological and molecular | bone marrow and/or splenic aspirate and blood | No information | Yes (S1 text) | No |
| Ostyn-2014 | Serological and/or parasitological | Unclear | No information |  | No |
| Cota-2014 | Serological and/or parasitological | Bone marrow | No information | No | No |
| Sundar-2014 | Serological and/or parasitological | bone marrow and/or splenic aspirate | No information | Not done | No |
| Mondal-2014 | Serological | Blood | No information | No | No |
| Jamil-2015 | Serological | Unclear | No information | No | No |
| Sundar-2015 | Parasitological | Unclear | No information | No | No |
| Goswami-2016 | Serological and/or parasitological | Unclear | No information | No | No |
| Wasunna-2016 | Parasitological | bone marrow or spleen or lymph node | Two slides per sample | No | No |
| Pandey-2016 | Serological and/or parasitological | Combination | No information | No | No |
| Borges-2017 | Serological and/or parasitological and/or molecular + culture | Combination | No information | No | PCR used in 4 patients for diagnosis |
| Rahman-2017 | Serological and/or parasitological | bone marrow and/or splenic aspirate | No information | No | No |
| Romero-2017 | Serological and/or parasitological + culture | Combination | No information | No | No |
| Alborzi-2017 | Serological and/or parasitological | Bone marrow | No information | No | No |
| Pandey-2017 | Serological and/or parasitological | bone marrow and/or splenic aspirate | No information | No | No |
| Kimutai-2017 | Serological and/or Parasitological and/or Clinical | Combination | No information | No | No |
| Mbui-2018 | Parasitological | bone marrow and/or splenic aspirate | No information | No | No |
| Goyal-2018 | Serological | Blood | No information | No | No |
| Sundar-2019 | Serological | Blood | No information | No | No |
| Diro-2019 | Parasitological | bone marrow and/or splenic aspirate | No information | No | No |
| Sinha-2019 | Parasitological | Spleen | No information | No | No |
| Goswami-2020 | Serological and/or parasitological | bone marrow and/or splenic aspirate | No information | No | No |
| Ekram-2021 | Parasitological | bone marrow and/or splenic aspirate | No information | No | No |

**Table 6: Methodology adopted for assessment of outcome at end of therapy and at the end of the study follow-up**

| **Author year** | **Test of cure assessment** | **End of study assessment** | **Tissue aspirates at test of cure** | **Tissue aspirates for confirmation of relapse** |
| --- | --- | --- | --- | --- |
| Gaeta-2000 | parasitological | clinical + parasitological | bone marrow | Aspiration not done |
| Sundar-2000c | clinical + parasitological | clinical + parasitological | spleen | bone marrow |
| Thakur-2000a | parasitological | clinical + parasitological | bone marrow/spleen | Done (unclear aspirate) |
| Sundar-2000b | clinical + parasitological | clinical | bone marrow/spleen | Unclear if aspiration done |
| Sundar-2000a | clinical + parasitological | clinical + parasitological | spleen | bone marrow |
| Thakur-2000b | clinical + parasitological | clinical + parasitological | bone marrow/spleen | Done (unclear aspirate) |
| Veeken-2000 | clinical | clinical + parasitological | lymph node | Done (unclear aspirate) |
| Villanueva-2000 | clinical + parasitological | clinical + parasitological | bone marrow | bone marrow |
| Thakur-2001a | clinical + parasitological | clinical + parasitological | spleen | spleen |
| Moore-2001 | clinical + parasitological | clinical and/or parasitological | spleen | Spleen |
| Ritmeijer-2001 | clinical and/or parasitological | clinical and/or parasitological | spleen | Done (unclear aspirate) |
| Thakur-2001b | clinical + parasitological | clinical + parasitological | spleen | Done (unclear aspirate) |
| Dietze-2001 | parasitological | clinical + parasitological | bone marrow/spleen | spleen |
| Sundar-2001 | clinical + parasitological | clinical + parasitological | spleen | spleen |
| Haidar-2001 | clinical + parasitological | Unclear | bone marrow/spleen | Monitored but not defined |
| Das-2001 | clinical + parasitological | clinical + parasitological | bone marrow/spleen | bone marrow |
| Sundar-2002 | parasitological | Unclear | Done (unclear aspirate) | Spleen |
| Sundar-2002a | clinical + parasitological | clinical + parasitological | bone marrow/spleen | bone marrow and/or spleen |
| Laguna-2003 | parasitological | clinical + parasitological | bone marrow/spleen/liver | bone marrow or tissue biopsy |
| Sundar-2003b | clinical + parasitological | clinical + parasitological | bone marrow/spleen | spleen |
| Figueras Nadal-2003 | clinical + parasitological | clinical + parasitological | bone marrow | Reported but not defined |
| Sundar-2003 | clinical + parasitological | clinical + parasitological | spleen | spleen |
| Syriopoulou-2003 | clinical + parasitological | Unclear | bone marrow | Bone marrow |
| Rijal-2003 | clinical + parasitological | clinical + parasitological | Done (unclear aspirate) | Done (unclear aspirate) |
| Thakur-2004a | clinical + parasitological | clinical + parasitological | spleen | spleen |
| Sundar-2004 | clinical + parasitological | clinical + parasitological | spleen | Done (unclear aspirate) |
| Thakur-2004b | clinical + parasitological | clinical + parasitological | bone marrow/spleen | Done (unclear aspirate) |
| Bhattacharya-2004 | clinical + parasitological | clinical + parasitological | Done (unclear aspirate) | spleen |
| Wasunna-2005 | parasitological | clinical + parasitological | spleen | spleen |
| Jha-2005 | parasitological | clinical + parasitological | Spleen | Spleen |
| Das-2005 | clinical + parasitological | clinical + parasitological | Done (unclear aspirate) | bone marrow and/or spleen |
| Ritmeijer-2006 | clinical/parasitological | clinical + parasitological | Spleen or lymph | Done (unclear aspirate) |
| Sundar-2006 | clinical + parasitological | clinical + parasitological | spleen | Spleen |
| Singh-2006 | parasitological | clinical + parasitological | Done (unclear aspirate) | spleen |
| Singh-2006 | parasitological | clinical + parasitological | Done (unclear aspirate) | spleen |
| Sundar-2007b | clinical + parasitological | clinical + parasitological | spleen | Done (unclear aspirate) |
| Sundar-2007a | clinical + parasitological | clinical + parasitological | Done (unclear aspirate) | bone marrow and/or spleen |
| Bhattacharya-2007 | parasitological | clinical + parasitological | spleen | spleen |
| Mueller-2007 | clinical + parasitological | clinical | Lymph node | Not actively assessed |
| Sundar-2008b | clinical + parasitological | clinical + parasitological | spleen | Done (unclear aspirate) |
| Sundar-2008a | clinical + parasitological | clinical + parasitological | spleen | Unclear if aspiration done |
| Thakur-2008 | clinical + parasitological | clinical + parasitological | Done (unclear aspirate) | bone marrow/spleen |
| Mueller-2008 | clinical + parasitological | clinical + parasitological | spleen | Mentioned but not defined |
| Sundar-2009a | parasitological | clinical + parasitological | spleen | Unclear if aspiration done |
| Das-2009 | clinical + parasitological | clinical + parasitological | bone marrow/spleen | Spleen |
| Sundar-2009b | clinical + parasitological | clinical + parasitological | spleen | spleen |
| Adam-2009 | No information | clinical | No information | No information |
| Shahian-2009 | No information | clinical | Unclear | Clinical suspicion only |
| Hailu-2010 | Not defined | clinical + parasitological | spleen/lymph/bone marrow | spleen/lymph/bone marrow |
| Thakur-2010 | clinical + parasitological | clinical + parasitological | spleen | spleen |
| Musa-2010 | parasitological | clinical + parasitological | Done (unclear aspirate) | Done (unclear aspirate) |
| Sundar-2010 | clinical + parasitological | clinical | spleen | Done (unclear aspirate) |
| Mondal-2010 | clinical | clinical + parasitological | Unclear | spleen |
| Rijal-2010 | clinical + parasitological | clinical + parasitological | Done (unclear aspirate) | Done (unclear aspirate) |
| Singh-2010 | parasitological | clinical + parasitological | Done (unclear aspirate) | bone marrow |
| Sinha-2010 | clinical + parasitological | clinical + parasitological | bone marrow/spleen | bone marrow/spleen |
| Sundar-2011b | clinical + parasitological | clinical + parasitological | bone marrow/spleen | Unclear if aspiration done |
| Sundar-2011c | parasitological | clinical | spleen | spleen and/or bone marrow |
| Sundar-2011a | clinical + parasitological | clinical + parasitological | spleen | spleen |
| Rahman-2011 | clinical + parasitological | clinical + parasitological | bone marrow/spleen | bone marrow/spleen |
| Sinha-2011 | clinical | clinical | Unclear | bone marrow/spleen |
| Sudarshan-2011 | parasitological | clinical + parasitological | peripheral blood | peripheral blood |
| Sundar-2012 | clinical + parasitological | clinical + parasitological | spleen | Spleen |
| Musa-2012 | parasitological | clinical + parasitological | spleen/bone marrow/lymph | bone marrow/lymph/spleen |
| Patra-2012 | parasitological | clinical + parasitological | Done (unclear aspirate) | bone marrow and/or spleen |
| Rijal-2013 | clinical + parasitological | clinical + parasitological + molecular | bone marrow | Done (unclear aspirate) |
| Khalil-2014 | parasitological | clinical + parasitological | Done (unclear aspirate) | bone marrow/lymph/spleen |
| Ostyn-2014 | clinical | clinical + parasitological | Unclear | Done (unclear aspirate) |
| Cota-2014 | clinical | clinical | Not carried out | bone marrow |
| Sundar-2014 | parasitological | clinical | bone marrow/spleen | Mentioned but not defined |
| Mondal-2014 | clinical | clinical + parasitological | Unclear | spleen |
| Jamil-2015 | clinical | clinical | Unclear | Mentioned but not defined |
| Sundar-2015 | clinical + parasitological | clinical + parasitological | bone marrow/spleen | spleen |
| Goswami-2016 | clinical + parasitological | clinical + parasitological | bone marrow/spleen | spleen |
| Wasunna-2016 | parasitological | clinical + parasitological | bone marrow/spleen/lymph node | Done (unclear aspirate) |
| Pandey-2016 | clinical | Unclear | Unclear | Mentioned but not defined |
| Borges-2017 | clinical | clinical | Unclear | Reported but not defined |
| Rahman-2017 | clinical + parasitological | clinical | bone marrow/spleen | Done (unclear aspirate) |
| Romero-2017 | clinical | clinical | Unclear | Clinical suspicion only |
| Alborzi-2017 | clinical | clinical + parasitological/serological | spleen | Done (unclear aspirate) |
| Pandey-2017 | clinical + parasitological | Unclear | bone marrow/spleen | spleen |
| Kimutai-2017 | clinical + parasitological | clinical + parasitological | Done (unclear aspirate) | Done (unclear aspirate) |
| Mbui-2018 | clinical + parasitological | clinical + parasitological | bone marrow/spleen | Reported but not defined |
| Goyal-2018 | No information | clinical + parasitological | Unclear if aspiration done | bone marrow and/or spleen |
| Sundar-2019 | clinical | clinical + parasitological | spleen | spleen |
| Diro-2019 | parasitological | Unclear | bone marrow/spleen | Assessed but not reported in this publication |
| Sinha-2019 | clinical + parasitological | clinical + parasitological | Done (unclear aspirate) | Spleen |
| Goswami-2020 | not defined | clinical + parasitological | Done (aspirates not clear) | Spleen |
| Ekram-2021 | clinical | clinical | Unclear | Mentioned but not defined |
